# Towards a complete characterization of common human polymorphic inversions and their functional effects

**DOI:** 10.64898/2026.08.03.26359593

**Authors:** Jon Lerga-Jaso, Elena Campoy, Marta Puig, lllya Yakymenko, Ruth Gómez- Graciani, Ricardo Moreira-Pinhal, Teresa Soos, Alba Vilella-Figuerola, Claudia Ramírez, Carla Giner-Delgado, Roser Zaurin, Sergi Villatoro, Alejandra Delprat, Marina Laplana, Mario Cáceres

**Author notes:** Equally contributing authors (Jon Lerga-Jaso, Elena Campoy, Marta Puig).

## Abstract

Structural variants (SVs) contribute substantially to genetic and phenotypic diversity, but their characterization is far from complete. Inversions are particularly interesting because they affect recombination and could have negative consequences on fertility. However, they are often missed due to their balanced nature, the repetitive sequences at their breakpoints and the fact that many are recurrent. Here, thanks to an in-depth analysis of >350 predictions from different studies, manual annotation, and accurate validation and genotyping in diverse populations, we have generated the largest and most reliable dataset of human polymorphic inversions to date, making it finally possible to determine their real functional and evolutionary impact. This unique resource totals 134 inversions, which in many cases consist of more complex rearrangements with additional insertions and deletions, and 61 inverted duplications that were used as a control. In particular, by rigorous imputation in available functional data, we have shown that many of the studied variants act as lead expression QTLs (eQTL) for different genes in multiple tissues. Moreover, several inversions are associated with epigenetic changes in chromatin accessibility, DNA methylation or histone marks. Finally, ∼20% of inversions are in high linkage disequilibrium (LD) with GWAS signals, including an inversion showing frequency differences across continents that is associated with body shape and height. Remarkably, when compared to SNPs, inversions tend to be enriched in functional effects, especially the largest ones that have already been proposed to act as supergenes, which could compensate for their potential fertility costs. Therefore, these findings highlight the important role that inversions can play in many organisms and reveal previously missing variants responsible for human phenotype variability.

## Introduction

New genomic techniques are revealing that the variation between individual genomes is much higher than previously thought. Structural variants (SVs) comprise different types of DNA sequence changes of more than 50 bp, that sometimes accumulate together in complex structural haplotypes (Porubsky and Eichler 2024; Collins and Talkowski 2025). Globally, SVs affect a larger fraction of the genome than single nucleotide polymorphisms (SNPs) and small indels (Collins et al. 2020; Byrska-Bishop et al. 2022; Ebert et al. 2021; Logsdon et al. 2025), suggesting that they could have a greater functional impact. Different large-scale projects have investigated the effect of these changes in gene expression (Chiang et al. 2017; Vialle et al. 2022; The GTEx Consortium 2017), phenotypic traits and disease susceptibility (Gudbjartsson et al. 2015; Bai et al. 2026; Gong et al. 2025). These studies have found that in general SVs have slightly more effects on gene expression than SNPs and could be involved in several phenotypes and diseases, although the number of new associations uncovered has been limited and few SVs act as lead variants). Part of the problem is that in most cases there is not a good definition of the SVs, with their precise location and accurate identification of the full set of individual variants. In addition, typically the number of available genotypes is relatively low and can include a high proportion of errors, which precludes reaching reliable conclusions.

Inversions are a particularly interesting type of SVs because, apart from the actual genome reorganization, they suppress recombination, which is a fundamental biological process, and, as a result, they can have negative effects on fertility, especially long inversions (Hoffmann and Rieseberg 2008; Puig et al. 2015a; Kirkpatrick 2010; Berdan et al. 2023). Accordingly, inversions of a considerable length that are maintained in populations probably compensate any fertility costs by positive or balancing selection (Gómez-Graciani et al. in prep.), and therefore they are more likely to have functional consequences than other variants. In fact, there is an increasing number of inversions associated with different phenotypic traits and adaptation in multiple organisms (Wellenreuther et al. 2025). Some good examples are the two well-known large inversions in 17q21.31 and 8p23.1 in humans, which have been related to many different effects and could act as supergenes (Stefansson et al. 2005; Salm et al. 2012; Campoy et al. 2022). Similarly, although the combination of different techniques is finally providing a complete picture of the whole catalogue of human SVs, only 298-456 inversions have been identified in the most recent studies (Collins et al. 2020; Byrska-Bishop et al. 2022; Ebert et al. 2021; Logsdon et al. 2025). This is several orders of magnitude less than that of other SVs and it supports the existence of a strong selection against inversions. However, due to the balanced nature and the breakpoint repeats of many inversions, they are still very challenging variants and just a small set of them have been analyzed in detail so far.

In the last years, there have been considerable efforts to validate and characterize precisely human polymorphic inversions, including the development of different PCR-based methods to genotype a subset of these variants in large samples (Pang et al. 2013; Martínez-Fundichely et al. 2014; Aguado et al. 2014; Vicente-Salvador et al. 2017; Giner-Delgado et al. 2019; Puig et al. 2020; Porubsky et al. 2022, 2023). In particular, highly accurate genotypes of 61 common inversions were generated in 95-550 individuals from diverse populations of the 1000 Genomes Project (1KGP), including 21 generated by non-homologous (NH) mechanisms and 40 mediated by non-allelic homologous recombination (NAHR) between inverted repeats (IRs) of 0.7-134 kb (Giner-Delgado et al. 2019; Puig et al. 2020). This has shown that NH inversions tend to have nearby tag SNPs in perfect linkage disequilibrium (LD), whereas most NAHR inversions occur recurrently, toggling back and forth in different individuals with a mutation rate ∼1000 times higher than that of SNPs (Giner-Delgado et al. 2019; Puig et al. 2020; Porubsky et al. 2022), which makes many of them very difficult to impute reliably. Nevertheless, despite the limited number of variants that could be analyzed, it is clear that inversions can have important effects. Apart from the two mentioned Chr. 17 and Chr. 8 inversions, several others have been associated with changes in expression levels or the rearrangement of the actual sequence of genes (González et al. 2014; Puig et al. 2015b; Giner-Delgado et al. 2019; Puig et al. 2020; González et al. 2020) and DNA methylation (Carreras-Gallo et al. 2022). Moreover, inversions have been associated with different phenotypic traits, such as obesity, diabetes, non-alcoholic pancreatitis, cognitive abilities or brain morphology (González et al. 2014; Rosendahl et al. 2018; Puig et al. 2020; Spracklen et al. 2020; Wang et al. 2023). Finally, human inversions with functional effects also show an enrichment of positive or balancing selection signals, supporting their potentially important evolutionary role (Giner-Delgado et al. 2019).

In this work, we extend the previous analyses to a more complete set of common human inversions (frequency >1-2.5%) that includes a significant fraction of those still missing, as well as additional resolved inverted duplications for comparison. This is done through an exhaustive characterization and validation of previous candidate inversions, plus the integration of the most recent 1KGP high-coverage (HC) data and a wide variety of functional datasets, achieving/making possible a more systematic and global quantification of the impact of these little-known variants. In addition, it also provides a much-needed benchmark of carefully resolved and annotated variants with reliable genotype information to facilitate the development of detection methods and future studies of SVs.

## Results and Discussion

### Inversion dataset generation

To ensure a representative view of human polymorphic inversions, the first step was to create the most complete and unbiased dataset of inversions possible, based on the available information. To do that we took advantage of the information available at the InvFEST database (https://invfestdb.researchmar.net/) when the project started (Martínez-Fundichely et al. 2014). Typically, inversion predictions have a considerably high error rate, especially those based on short reads and paired-end mapping (PEM) (Martínez-Fundichely et al. 2014; Aguado et al. 2014; Vicente-Salvador et al. 2017). Therefore, we first tried to validate and define the exact breakpoints of the maximum number of inversions that have been identified more or less accurately using different methods. Specifically, targeted candidates included previously validated inversions of >50 bp from PEM data (Korbel et al. 2007; Hehir-Kwa et al. 2016), which included a detailed analysis of fosmid PEM predictions (Kidd et al. 2008, 2010; Martínez-Fundichely et al. 2014), or with >1% global frequency in the 1KGP (Sudmant et al. 2015) and all those obtained from long reads, which map more reliably (Chaisson et al. 2015; Huddleston et al. 2017) (Table 1). Those inversions located entirely within a repetitive element or with breakpoints in complex segmental duplications (SDs) were not considered (Table S1).

**Table 1.**
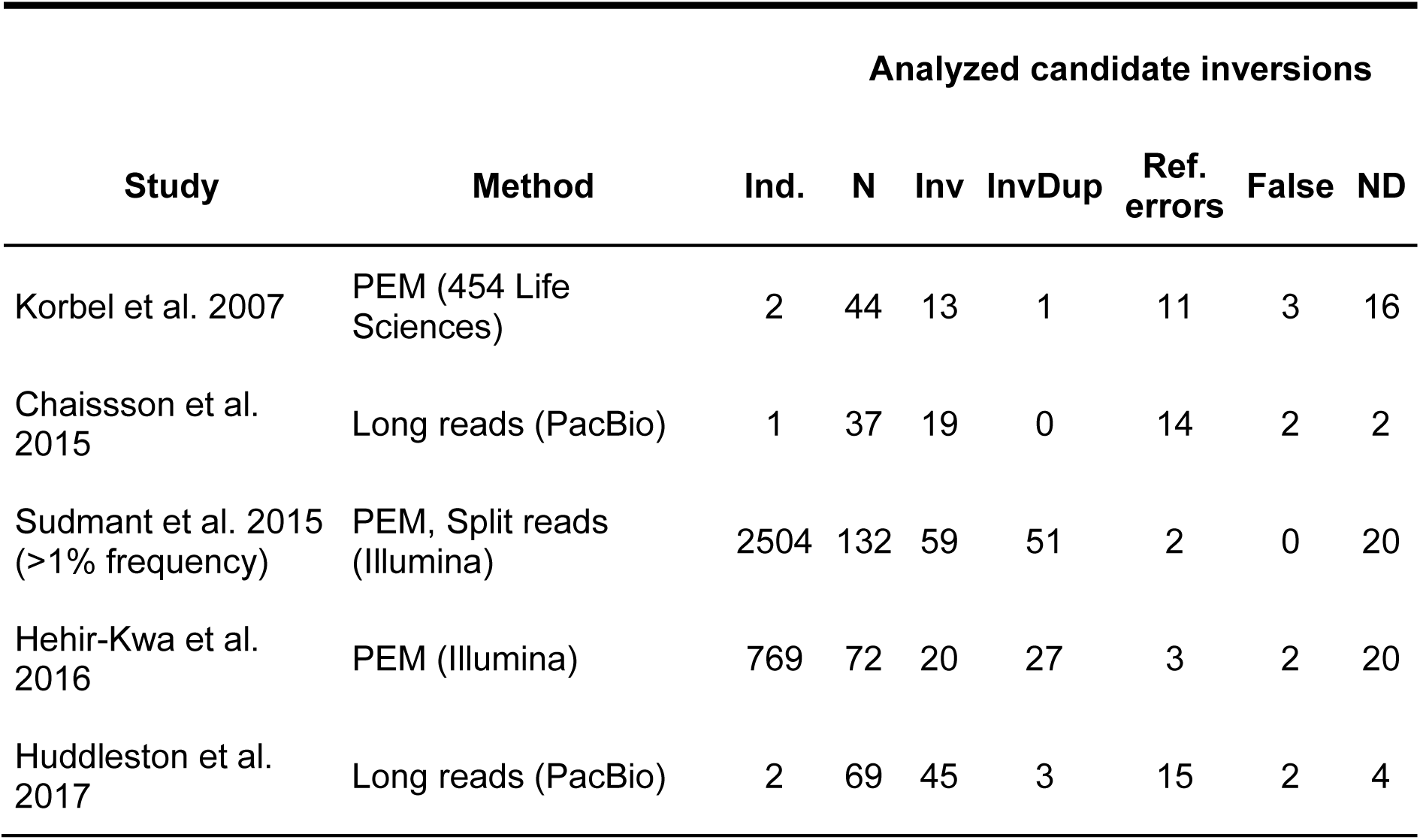
Summary of the validation results of the candidate inversions from the different studies analyzed in this work. N indicates the number of independent inversions identified in each study after merging redundant predictions. Ref. errors are regions in the corresponding human genome reference sequence that have been validated as assembled incorrectly in the inverted orientation (Vicente-Salvador et al. 2017). ND corresponds to regions for which no sequence in the alternative orientation supporting the inversion could be found or that they were too complex to be reliably analyzed. PEM, paired-end mapping.

To confirm the presence of these putative inversions and identify sequences with the inverted allele to define precisely their breakpoints and other possible SVs involved, we did an exhaustive search of available sequences with the alternative orientation by BLAST in both the NCBI core non-redundant nucleotide sequence database and 69 available genome sequence assemblies from 35 individuals (Table S2; see Methods for further details). This was complemented by the generation of our own sequences in a few specific cases (Table S1). Next, the specific SVs and their breakpoints were resolved by the manual analysis of the BLAST results and genome sequence annotation (Table S1). However, it is difficult to assemble reliably the sequence of many NAHR inversions mediated by IRs of >90% identity, especially for long repeats. In addition, in some cases available alternative sequences might contain errors and not be totally representative of the alternative allele. Therefore, to make sure that they correspond to real inversions, both breakpoints of all those candidates supported by alternative sequences or mediated by IRs for which PCR assays could be designed were validated experimentally in a small set of samples by regular PCR or by inverse PCR (iPCR), which made possible to interrogate inversions with IRs of ∼1-25 kb at their breakpoints (Aguado et al. 2014) (Table S1).

In total, from the five different studies together, we analyzed 354 candidate inversions, and the sequence analysis allowed us to resolve 292 of them (82.5%), including 98 potential inversions and 65 InvDups (Table 1). Of those, 45 of the resolved predictions corresponded to 22 already known inversions and 3 possible assembly errors in the human genome reference sequence, in which all the analyzed sequences show the alternative orientation (Vicente-Salvador et al. 2017). Finally, another 9 inversion predictions were invalidated due to different types of sequence mapping problems (Table S1). Therefore, excluding the inversions already characterized in previous studies (Aguado et al. 2014; Vicente-Salvador et al. 2017; Giner-Delgado et al. 2019; Puig et al. 2020), it was possible to define accurately the breakpoints and mechanism of generation of 66 new inversions and 65 InvDups, which practically triplicates those previously characterized (Table S3). This included the precise annotation of multiple additional rearrangements associated with these variants, which is critical for their correct genotyping and follow-up analysis. Of those new variants, we tested by PCR or iPCR the 66 inversions and 5 InvDups and all of them were validated. With regard to the different studies, they all showed a good validation of the resolved variants, ranging from 89.3% (Korbel et al. 2007) to 100% (Sudmant et al. 2015), although in some cases this comprised many human reference genome assembly errors (1.8-39.3%) and approximately half of those in Sudmant et al. (2015) and Hehir-Kwa et al. (2016) were InvDups instead of inversions (Table 1).

### Inversion genotyping in human populations

As already mentioned, to determine the functional and evolutionary impact of these variants it is crucial to have reliable genotypes in large population samples. Therefore, we tried to genotype in as many individuals as possible all inversions and InvDups that had been validated by sequence or PCR in this and previous studies and for which there is no genotype data in different human populations (Table S3). This was done by two complementary approaches, PCR-based methods and bioinformatic identification of available sequence reads supporting both alleles of those variants without repetitive sequences at the breakpoints using the program BreakSeq (Lam et al. 2010; Lucas-Lledó et al. 2014). This strategy is summarized in Figure 1.

**Figure 1.**
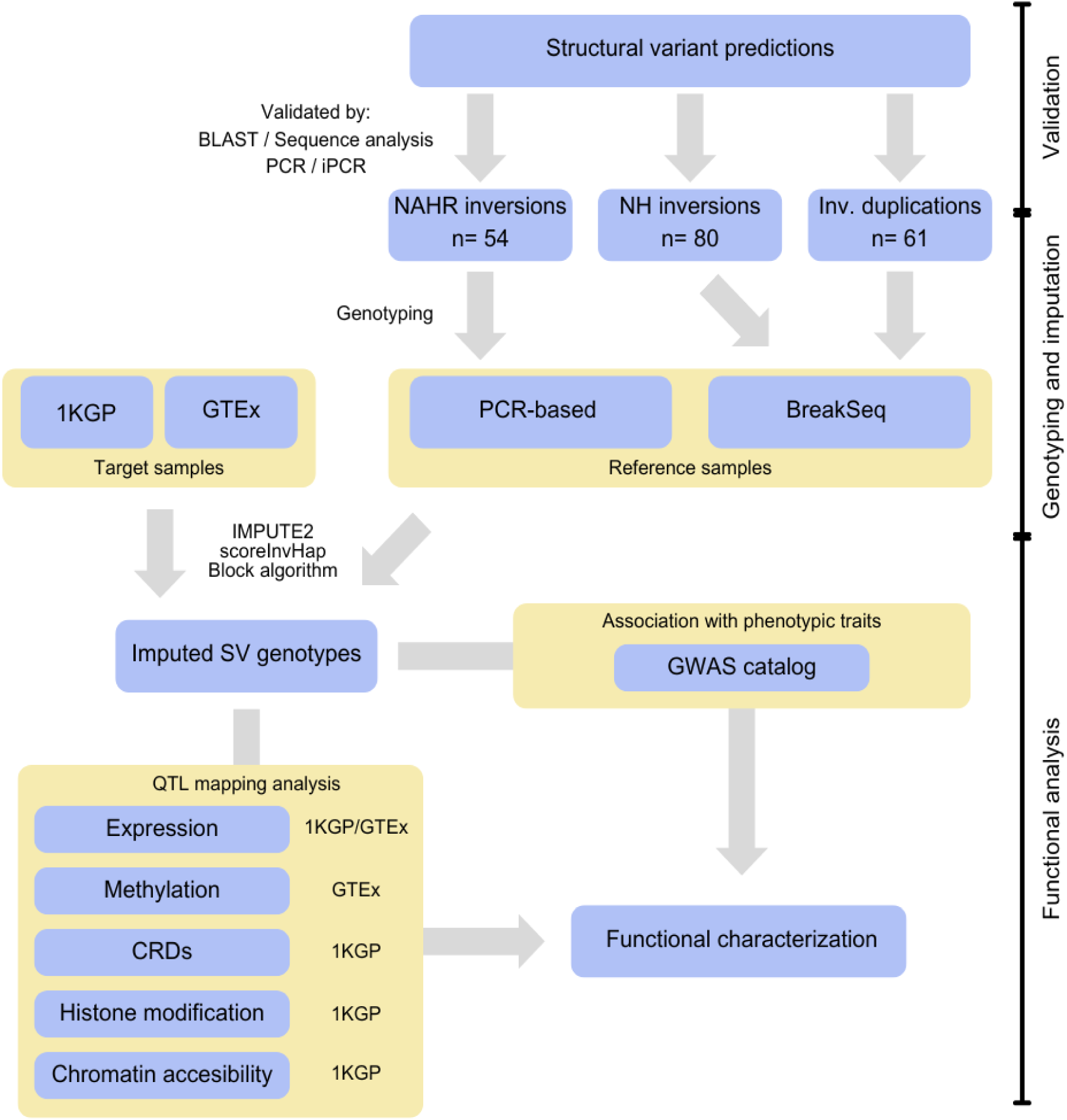
General diagram of the strategy used in this study to generate the large-scale dataset of inversions and related variants and to determine their functional effects using different types of available data.

Specifically, one breakpoint of 74 experimentally validated inversions, consisting on the 66 new inversions described here and 8 inversions annotated and validated in previous studies (Aguado et al. 2014; Vicente-Salvador et al. 2017) plus 2 associated deletions, were genotyped experimentally by PCR (64) or iPCR (12) in 95 individuals of YRI, CEU and ASN ancestries as a representation of human global diversity (Puig et al. 2020) (Table S3). This allowed us to do a preliminary analysis of LD with other variants across human populations and validate BreakSeq genotypes (see below). Moreover, for 13 NAHR inversions without perfect tag SNPs (*r*^2^ = 1) in the previous global diversity samples or mediated by IRs of >1 kb, the PCR genotyping was extended to 136 additional individuals of CEU, TSI and YRI ancestry (Table S3), which encompassed the maximum number of available samples with gene expression data (Lappalainen et al. 2013; Giner-Delgado et al. 2019). The larger genotyping sample for NAHR inversions compensates in part the lack of BreakSeq genotypes (see below) and helps to improve imputation in other individuals and functional analysis of potentially recurrent inversions. These genotypes provided also extra information of inversions previously analyzed only in the CEU population (HsInv0286 and HsInv1122) (Aguado et al. 2014; Vicente-Salvador et al. 2017) or in the 95 initial individuals by the ddPCR technique (Hsinv1057) (Puig et al. 2020). Finally, for several of the variants, between 1 and 326 additional genotypes were obtained for certain populations or samples to get a better idea of their actual prevalence and population distribution, for example in the case of those with low frequency (Table S3).

In parallel, we took advantage of the available sequence reads from the 3202 1KGP-HC individuals (Byrska-Bishop et al. 2022) to obtain information of all the inversions originated by NH mechanisms with manually curated breakpoints lacking highly identical repeats from this and previous studies (79) in a much larger and worldwide sample. These variants were genotyped by a considerably improved version of BreakSeq (Lam et al. 2010; Lucas-Lledó et al. 2014) using a variable number of pairs of 300 bp sequence probes specific to the breakpoints of each orientation, which for a typical inversion consisted of a total of four probes (two for each breakpoint), but could range from just two to up to eight different probes (see Methods). In addition, given the good performance of BreakSeq genotyping (see below), the analysis was extended to 61 of the 65 sequence-resolved InvDups in which specific sequence probes could be designed, in order to compare them to the real inversions (Table S3). This also allowed us to interrogate different possible independent SVs (such as the deletion affecting the HsInv1122 inversion, genotyped by PCR in a much smaller sample set, and two other deletions in HsInv1306 and HsInv1141) and have a better coverage of more complex rearrangements (e.g. HsInv1065).

Using BreakSeq, we obtained reliable results for 99.5% of the total possible genotypes, with more than 99.3% of the samples genotyped for 132 of the 143 analyzed variants, including all the inversions. We confirmed the robustness and accuracy of this methodology by comparing the BreakSeq genotypes and those generated by PCR-based techniques for 77 inversions and a deletion in 87 to 523 individuals from this and previous studies (Vicente-Salvador et al. 2017; Giner-Delgado et al. 2019), which showed only two discrepant genotypes for HsInv0113 with no clear cause. Thus, BreakSeq worked quite well for most of the variants. The only exceptions were six InvDups, in which more strict genotyping criteria were applied, due to a lack of differentiation of homozygotes and heterozygotes caused probably by some probe mapping problems (see Methods), resulting in discarding a high proportion of unreliable genotypes (7.6-49.3%). In any case, this analysis made it possible to expand considerably the information available for a large fraction of inversions and InvDups. In addition, the BreakSeq data showed that HsInv1868 is likely a sequence error or a new mutation of the CHM13 cell line used for the T2T reference genome sequence, since despite having relatively simple breakpoints, it was not found in two other CHM13 assemblies or in any of the 3202 1KGP samples. Similarly, it confirmed the only one of the potential hg38 assembly errors not mediated by IRs that could be tested (HsInv1874), because all the samples were homozygous for the *O2* orientation. Lastly, the genotypes of the two new possibly independent deletions matched always those of the nearby HsInv1306 inversion or HsInv1141 InvDup, indicating that they form part of a larger rearrangement.

Finally, all the data generated was merged with the previous experimental genotypes of 61 inversions in many of the same 1KGP samples, ranging from the 95 YRI, CEU and ASN individuals of Puig et al. (2020) to the full set of 551 individuals of Giner-Delgado et al. (2019), plus 40-121 additional available genotypes of the 17q21.31 and 8p23.1 inversions obtained by FISH (Antonacci et al. 2009; Steinberg et al. 2012; Salm et al. 2012) (Table S3). Therefore, this represents the largest population dataset of highly reliable information from human inversions, totaling 36,269 genotypes obtained by PCR-based methods and FISH (with 95- 557 samples per inversion) and 442,990 genotypes by BreakSeq (with 1624-3202 samples per variant) of 197 inversion related variants (54 NAHR inversions, 80 NH inversions, 61 InvDups and 2 independent deletions not completely linked with the previous variants) (Figure 2). Although functional and evolutionary studies of part of the inversions had already been carried out (Puig et al. 2015b; Giner-Delgado et al. 2019; Puig et al. 2020; Campoy et al. 2022), they were included to achieve a more global and complete characterization of the functional impact of these little studied variants with the newest data and analysis methods.

**Figure 2.**
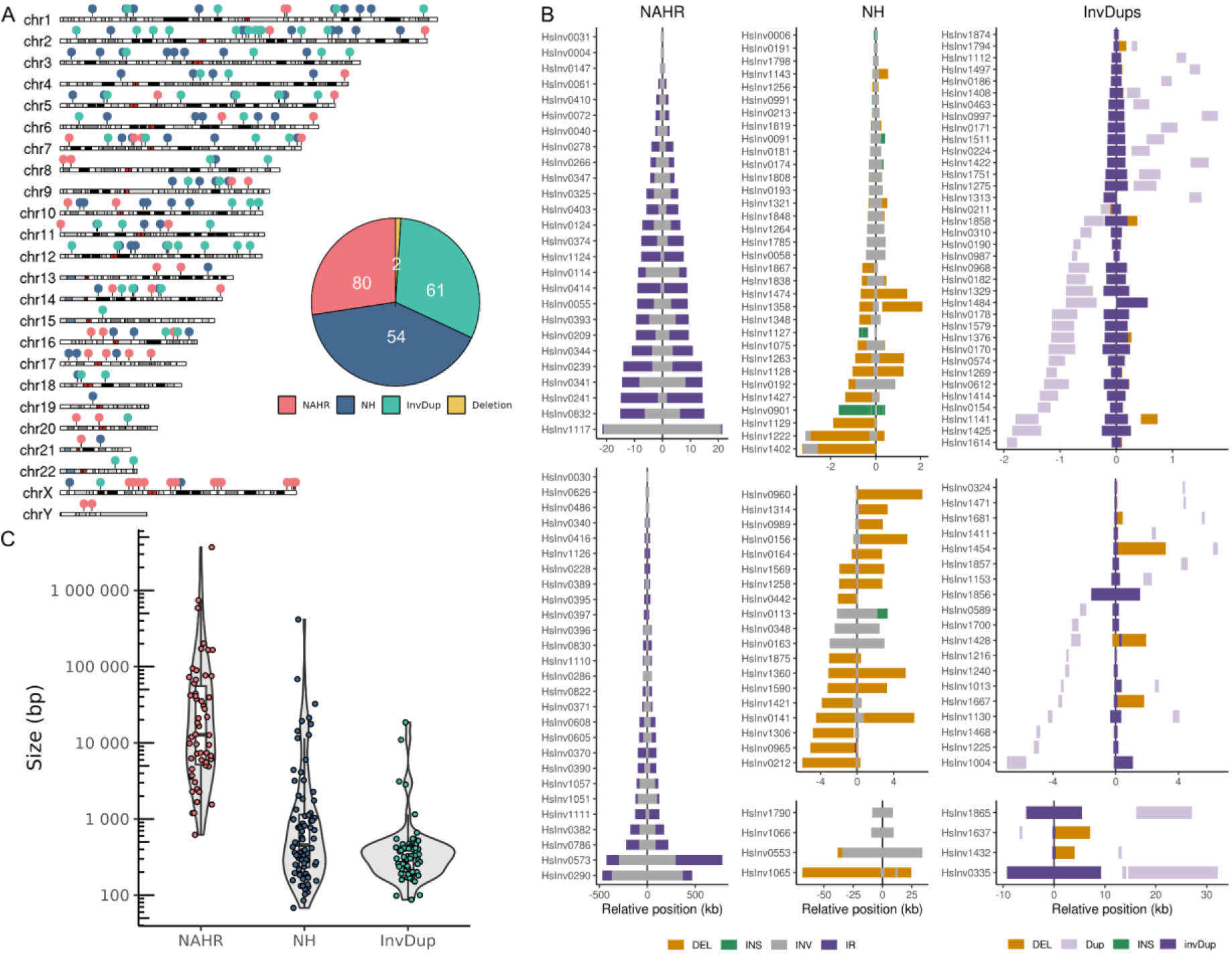
Summary of the inversion-related variants dataset. **A.** Distribution of validated inversions, inverted duplications (InvDup) and two additional independent deletions in the human genome. Inversions are divided in those generated by non-homologous mechanisms (NH) or non-allelic homologous recombination (NAHR). **B.** Representation of the main characteristics of the inversions and InvDups studied. The size of the inverted region of both NH and NAHR inversions is represented in gray, flanking inverted repeats (IRs) of NAHR inversions in purple and other deletions (Del) and insertions (Ins) in the derived allele associated with NH inversions and InvDups in orange or green, respectively. For InvDups, the original fragment that is duplicated is shown in light purple and the copy inserted in reverse orientation in dark purple. **C.** Size distribution of NAHR and NH inversions and InvDups showing significant differences among them (Kruskal-Wallis test of the three groups: *P* < 2.2 x 10^-16^. Dunn pairwise test: NAHR vs. NH inversions, *P* < 1 x 10^-5^; NH inversions vs. InvDups, *P* = 0.0165; NAHR inversions vs InvDups, *P* < 1 x 10^-5^).

### Human inversion catalogue

The current catalogue, generated by the aggregation of inversions detected and validated using different techniques, is likely a good representation of common human polymorphic inversions diversity, missing mainly some known inversions with large and complex IRs at the breakpoints (Martínez-Fundichely et al. 2014), which could not be genotyped using the available methods, or low frequency candidates (∼1-5% frequency) without supporting sequences (Table S1).

Specifically, the 134 inversions, plus the 61 InvDups and 2 independent inversion-associated deletions, were well distributed across the genome (177 in autosomes, 18 in Chr. X and 2 in Chr. Y) (Figure 2A). Moreover, thanks to the manually-curated precise breakpoint annotations, the mechanism of generation of all the variants could be defined (Vicente-Salvador et al. 2017; Giner-Delgado et al. 2019). As already mentioned, 54 of the inversions were likely generated by NAHR mechanisms between highly identical IR blocks of 140 bp to 1.2 Mb, consisting of SDs, TEs or unique sequences, whereas the 80 remaining inversions, all the InvDups and the 2 deletions were generated by NH mechanisms (Figure 2B; Table S3). Importantly, the majority of NH variants contain other small rearrangements together with the inversions or InvDups, consisting mainly of deletions and insertions of duplicated sequences from nearby regions or even other chromosomes (Figure 2B; Table S3). Therefore, some of the rearranged sequences can be quite complex, including for example 12 deletions, an insertion and an inversion in HsInv0901, or 3 deletions, 2 insertions and 2 inversions in HsInv1065. In addition, in most of the inversions and InvDups including additional rearrangements, pairs of microhomology sequences can be found between the breakpoints of the different changes, strongly indicating that they were generated in a single fork stalling template switching (FoSTeS) or microhomology-mediated break induced replication (MMBIR) event (Table S3) and supporting the major role of this process in NH inversions and InvDups origin. In a few inversions, InvDups and a deletion without other rearrangements, there was also microhomology at the breakpoints, suggesting that they could be generated by other simple repair mechanisms like microhomology-mediated end joining (MMEJ), whereas the rest of the variants originated apparently by non-homologous end joining (NHEJ) and two InvDups by retroposition (Table S3).

Regarding the size of the actual inversions, they range from 68 bp to ∼4 Mb. However, most of them are relatively small (median = 1.7 kb; mean = 55.6 kb), with 96 (71.6%) being shorter than 10 kb and only 7 (5.2%) exceeding 100 kb. As previously described (Giner-Delgado et al. 2019; Puig et al. 2020), NAHR inversions (median = 12.3 kb; mean = 125.7 kb) tend to be clearly larger than NH inversions (median = 0.5 kb; mean = 8.4 kb) (Figure 2C). In the case of InvDups, the size distribution looks quite similar to that of NH inversions, but they actually tend to be slightly smaller (mean = 0.9 kb) (Figure 2C). When compared to the main other largest dataset focused uniquely on inversion discovery (Porubsky et al. 2022), the size of inversion-like variants in our catalog was significantly smaller (mean 38.2 kb *vs* 245 kb; Mann-Whitney U test, *P* < 2.2 x 10^-16^). These discrepancies likely result from differences in the detection techniques employed, with long reads and Strand-seq playing a much bigger role in Porubsky et al. (2022), which allows the identification of larger inversions mediated by complex repeat blocks that are missing in our study, whereas we include a higher proportion of smaller variants below the Strand-seq detection limit. Additionally, the size distribution of InvDups also differs markedly between the two datasets, with Porubsky et al. (2022) reporting substantially larger events. However, the precision in the definition of these changes may also play a role in the observed differences.

### Inversion frequency and distribution in human populations

The diverse inversion dataset and accurate genotype information, including in many cases a large number of samples of multiple populations, enables for the first time an unbiased analysis of inversion frequency and distribution in humans and a preliminary assessment of the possible action of selection. To do that, we first used different types of evidence available to determine the ancestral orientation for the 61 NH inversions and 18 of the 54 NAHR inversions, the remaining being recurrent between species or lacking reliable information (Giner-Delgado et al. 2019; Diaz-Ros et al. 2026) (Table S4). As expected, in the large majority of the inversions the orientation present in the hg38 reference genome (*O1*) was the ancestral and the alternative orientation (*O2*) was the derived one (Giner-Delgado et al. 2019). Similarly, in all the InvDups except one (HsInv1874), hg38 included the ancestral non-duplicated allele (Table S4).

Next, based on the unrelated samples from the 1KGP-HC dataset for most NH variants and the experimental genotypes of the rest of inversions, the derived allele frequency (DAF), when the ancestral allele was known, or the minor allele frequency (MAF) otherwise was calculated for all variants globally across all individuals and for the five main superpopulations: African (AFR), European (EUR), South Asian (SAS), East Asian (EAS) and American (AMR) (Table S4). Except for one singleton inversion (HsInv0486, found exclusively in NA15510), the global frequency of the variants ranged from 0.02% (HsInv0335) to 87.6% (HsInv0004) (Table S4), and, as expected, 97.0% of the variants had a frequency >1%. This includes several low-frequency variants that are only polymorphic in one superpopulation, half of which were analyzed here for the first time: HsInv0097, HsInv0192, HsInv1051, HsInv1057, HsInv1471, HsInv1808 and HsInv1878 in AFR, HsInv0335 in EAS, and HsInv1110 in EUR. To investigate whether the different types of inversion-like variants are more or less frequent than expected under neutrality, we compared their frequency distribution to a null distribution derived from neutral SNPs (see Methods). Both NH inversions and InvDups showed a similar frequency across populations, which did not differ significantly from that of random SNPs (Figure S1). However, consistent with previous results, frequency of NAHR inversions was higher than expected in all populations except EAS (Figure S1), probably due to recurrence driving their frequency towards intermediate values, with one third of NAHR inversions (29.6%) showing a global MAF between 0.4 and 0.5.

To identify potential signals of natural selection in specific populations, we examined frequency differences across continents in our dataset using the fixation index (*F_ST_*) and compared it with that of neutral SNPs (see Methods). The average *F_ST_* considering all the populations together for inversion-like variants in autosomes or Chr. X was 0.080 and 0.129, respectively, which is similar to with that of SNPs. Nevertheless, three NH inversions (HsInv1402, HsInv1421 and HsInv1790), one NAHR inversion (HsInv0389) and two InvDups (HsInv1275 and HsInv1468) were in the top 1% of *F_ST_* values (Figure 3), with 30 additional variants in the top 5% (Table S5). These results coincide well with those of the 61 inversions already analyzed, with significant *F_ST_* differences previously identified for many of the inversions in common (Giner-Delgado et al. 2019; Puig et al. 2020). Moreover, we also identified 11 variants with extremely low global population differentiation, although all of them were present at low frequencies.

**Figure 3.**
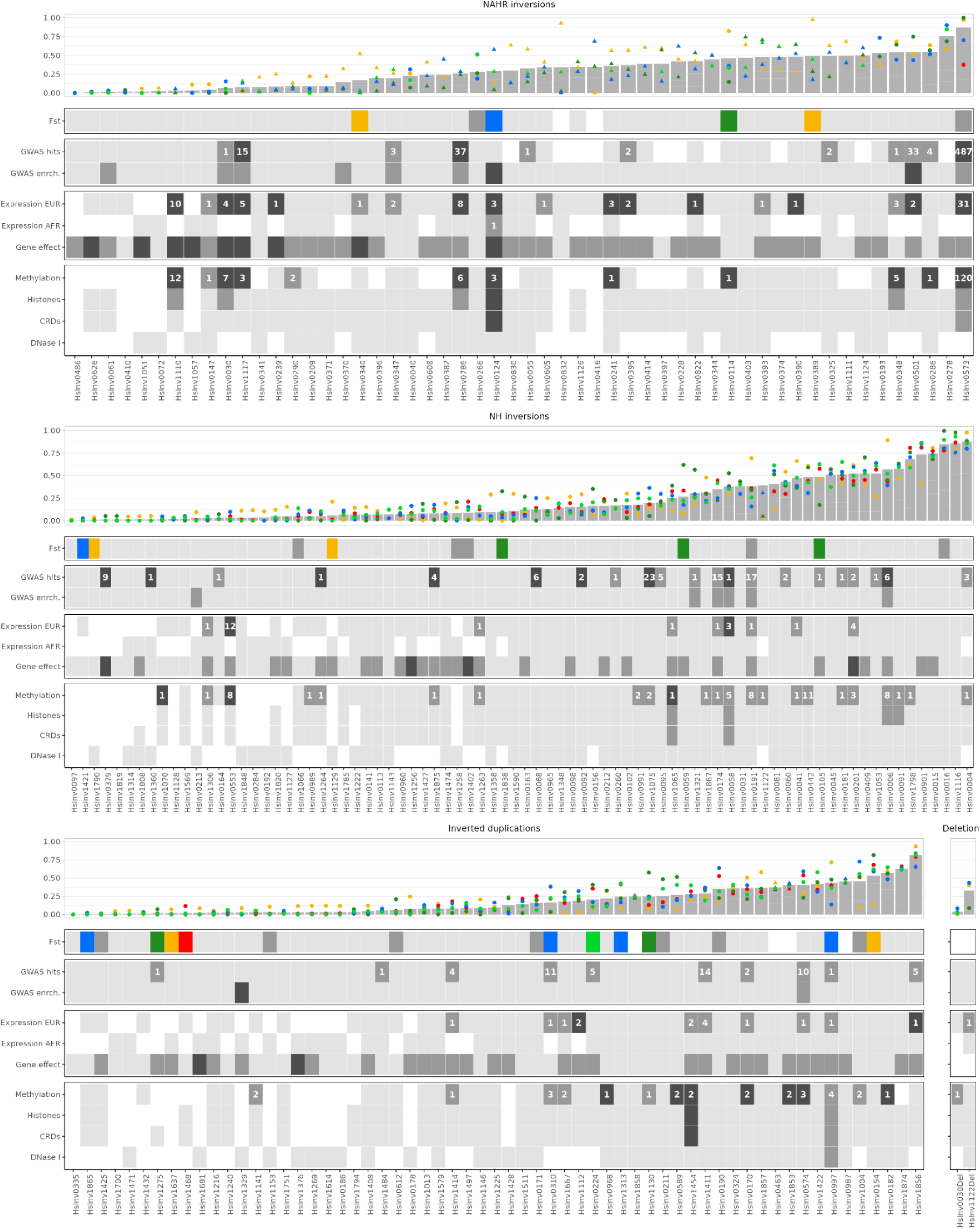
Summary of the functional and evolutionary information of all the variants analyzed in this study. For the different variant types, the top graph shows the frequencies in the available unrelated individuals from the five main super-populations (represented in different colors) of the minor (MAF) or derived (DAF) allele as triangles or dots, respectively, and the gray bar indicates the corresponding global frequency. The rectangular graphs below summarize the results of the different analysis performed as squares on a gray scale, with white meaning that the test could not be performed, light gray that no significant findings were found, and dark gray and black increasingly significant results according to the *P* values of the different tests. For the *F_ST_* analysis, dark gray corresponds to a *F_ST_* value across all populations in the top 5% of the SNP distribution and the colors indicate when there is a specific super- population that is significantly different than the others. In the GWAS-hits analysis, the LD between the variant and the SNP previously associated to the trait is represented in dark gray (*r*^2^ > 0.8) and black (*r*^2^ = 1), whereas the numbers within the boxes correspond to the amount of GWAS associations with each variant. In the gene expression and methylation analyses, the number indicates the total genes or cytosines with expression or methylation changes in at least one tissue significantly associated with each variant. In the gene effect, black highlights the cases where a variant disrupts a gene, transcript or exon, whereas dark grey those in which the variant overlap genes, is intronic or exchanges genic sequences.

### Inversion association with other variants and imputation

LD with neighboring variants provides information about the origin of the inversions and makes it possible to impute them in additional samples. We used the 1KGP high coverage accurate information to search for associations between 192 inversion-like variants (excluding two located in Chr. Y and a singleton inversion) and SNPs (including also small indels) located within the inversion or 500 kb flanking sequences at both sides. We considered perfect tag SNPs those with an LD value of *r*^2^ = 1 for the inversions genotyped by PCR-based methods (84-489 individuals) and *r*^2^ ≥ 0.975 for the variants genotyped with BreakSeq (1298-2593 individuals), to allow for low frequency recombination events or genotype errors in either the SNPs or the inversion-like variants. In total, we found that 117 of the analyzed variants (60.9%) have between 1 and 2542 global tag SNPs across all populations analyzed (Table S6). However, this number is higher in NH inversions (73/80, 91.3%) and InvDups (39/61, 63.9%), whereas only 9.8% (5/51) of the NAHR inversions have tag SNPs. This difference had been previously reported and it was related to a high level of recurrence of repeat-mediated inversions, which can occur several times on diverse sequence backgrounds (Giner-Delgado et al. 2019; Puig et al. 2020; Porubsky et al. 2022). In addition, inversions may be more likely to present tag SNPs than InvDups due to the suppression of recombination in heterozygotes, which promotes the differentiation of inverted and non-inverted haplotypes.

When we compared the LD for 59 inversions analyzed in previous studies using a lower number of genotypes for many of them (∼500 vs. ∼2500) and a slightly different set of SNPs (1KGP Phase 3 vs. high coverage) (Giner-Delgado et al. 2019; Puig et al. 2020), results were highly consistent. Of the 23 inversions that had previously perfect tag SNPs, 22 still maintained the tag SNPs in the new dataset, which were mostly the same, indicating a very robust association, and just one inversion lost its only tag SNP (Table S6). As expected, 15 of those inversions had less perfectly tagging SNPs when more genotypes were analyzed, contributing to eliminate spurious SNPs in high LD due to a small number of samples. However, another six inversions acquired new tag SNPs, some of which were probably not present in the first SNP set, including HsInv0105 that did not have any tag SNPs before (Giner-Delgado et al. 2019). Therefore, the strong LD with nearby SNPs for many of the analyzed variants based in large sample sizes indicates that they can be confidently used as proxies for the variant alleles.

Next, we used the linked SNP information to determine the status of inversion-like variants in other 1KGP or GTEx samples with relevant functional data and no PCR or BreakSeq genotypes (Table S7). If present in the corresponding dataset, genotypes were determined based on the identified tag SNPs. Otherwise, we used IMPUTE2 (Howie et al. 2009) to infer the genotype of those variants showing a good imputation accuracy (*r*^2^ ≥ 0.8, see Methods). This resulted in notably different genotype obtention patterns for the diverse datasets depending on the origin of the variants (Table S7). All polymorphic NH inversions and InvDups could be genotyped in 1KGP AFR and EUR samples using the available BreakSeq and tag SNP information (Figure 4). Conversely, because of the lack of BreakSeq data and tag SNPs, NAHR inversions were predominantly inferred with IMPUTE2, although around one third of them could not be imputed accurately (Figure 4), due likely to recurrence and/or low frequency (Yakymenko et al. 2026). In addition, for two of the largest inversions that could not be imputed with IMPUTE2, we used scoreInvHap (Ruiz-Arenas et al. 2019) (Table S7), which had previously shown good imputation results for the 8p23.1 inversion in EUR (Campoy et al. 2022).

**Figure 4.**
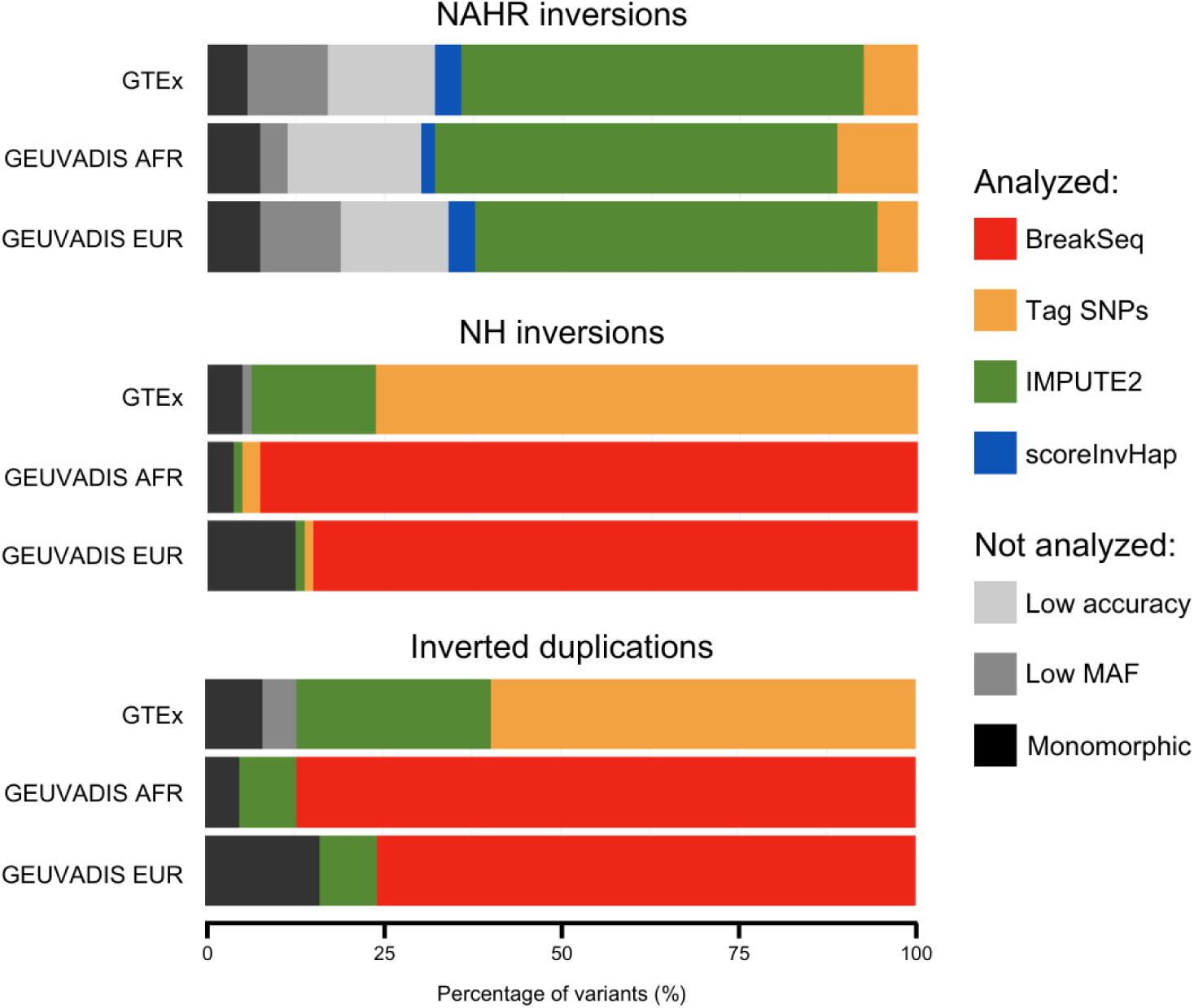
Distribution of variants determined using different methods in the 1KGP AFR and EUR and GTEx datasets. Variant genotypes were inferred using BreakSeq, tag SNPs, IMPUTE2, or scoreInvHap. Some variants were excluded from the analysis (not analyzed) because they were monomorphic in the studied population or no alternative alleles were imputed due to low MAF in the reference panel or they showed low imputation accuracy.

Specifically, in the 1KGP dataset, out of the 192 inversions and InvDups analyzed, 163 (84.9%) could be genotyped in the AFR superpopulation (126 by BreakSeq, 8 by tag variants, 28 by IMPUTE2, and 1 by scoreInvHap) and 150 (78.1%) in the EUR superpopulation (113 by BreakSeq, 5 by tag SNPs, 31 by IMPUTE2 and 1 by scoreInvHap) (Figure 4; Table S7). The remaining variants were supposedly monomorphic in each population (11 in AFR, 28 in EUR) or the IMPUTE2 or scoreInvHap imputation accuracy was below the threshold (18 in AFR, 14 in EUR), and they were excluded from further analyses. On the other hand, since there is no BreakSeq data, in the EUR samples from GTEx, genotypes of 158 (82.3%) inversions and InvDups were inferred using tag SNPs (101), including 12 low frequency variants that were monomorphic in 1KGP EUR, or imputation (56 by IMPUTE2 and 1 by scoreInvHap), that was extended to several cases in which the tag SNPs were not present in the GTEX sequence variation data (Figure 4; Table S7). The rest of variants were monomorphic (20) or not imputable (14). Lastly, except for HsInv0030 deletion in AFR, the genotypes of the two inversion associated deletions could also be accurately determined in the three datasets using BreakSeq and tag SNPs or IMPUTE2 (Table S7). Thus, combining different data sources allowed us to obtain reliable information for the maximum number of inversion-related variants in large numbers of samples, but still a significant proportion of NAHR inversions could not be analyzed (Figure 4).

### Impact of inversion-like variants on genes

Previous studies have shown that inversions can significantly impact genes (Chiang et al. 2017; Giner-Delgado et al. 2019; Puig et al. 2020). Thus, thanks to this new comprehensive and accurate dataset of human polymorphic inversions and related variants, we were able to extend our knowledge of inversion functional consequences. As a first step, we determined the overlap of the characterized variants with annotated genes and transcripts from GENCODE (Mudge et al. 2025) and classified them into different groups based on the potential consequences on the genes (Figure 5A). Nearly half of the 195 inversions and InvDups analyzed (47.2%, 92/195) are intergenic, while 103 showed some overlap with coding or non-coding genes, with 60 located just within introns and 43 including exons (Table S8). As expected by the larger size of the inverted region and IRs, NAHR inversions accounted for 81.4% (35/43) of the variants overlapping exons, while the great majority of NH inversions and InvDups are intronic or intergenic (Figure 5B). Among the exon-overlapping inversions, most simply invert completely genes contained within them (20) or overlap partially with a gene (8), with no clear consequences. However, there were also more severe cases of inversions disrupting a gene (11), inverting one exon (1) or causing a sequence exchange between different genes (3), several of which have already been described before (Giner-Delgado et al. 2019). Of those not previously analyzed, one of the most severe effects is the disruption of protein-coding transcripts, as observed in HsInv0626 (*VIPR2*), HsInv1117 (*AKR1C2*) and HsInv1402 (*RASGRP3*), whereas HsInv1057 results in an exon exchange between *TCP10* and *TCP10L2* located at the inversion breakpoints. Conversely, none of the inverted duplications disrupted any protein-coding genes and only two disrupted pseudogenes. Finally, the proximity of intergenic variants to nearby genes may also affect regulatory regions and 2 NAHR inversions, 7 NH inversions and 4 inverted duplications) are located within 5 kb of a protein coding or long non-coding gene.

**Figure 5.**
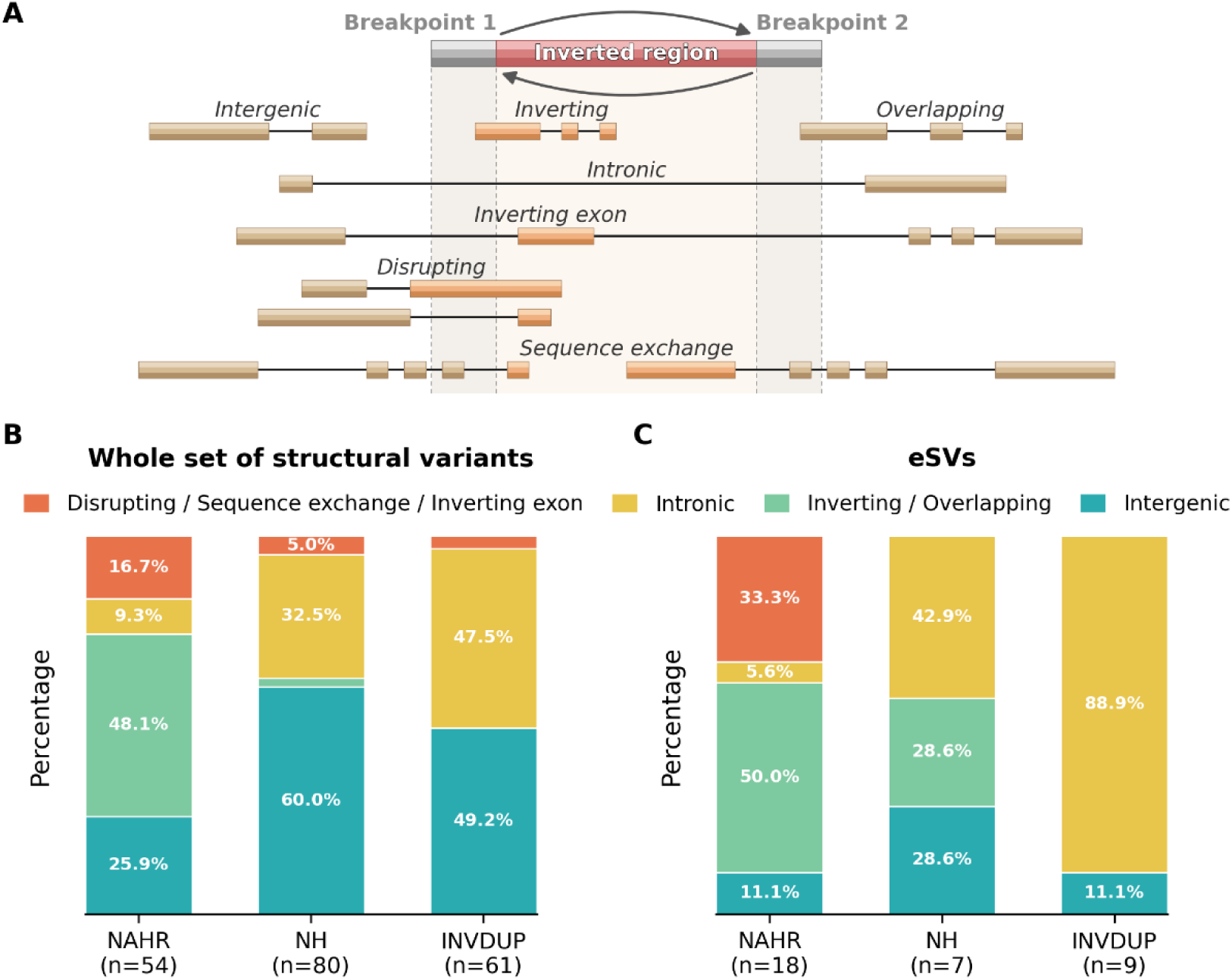
Mutational effects of inversions and inverted duplications. **A.** Classification categories for the SVs based on their position relative to gene structure. **B.** Proportion of each functional impact category for each type of inversion-like variant, considering the whole set of structural variants. **C.** Same proportions but considering only SVs associated with gene expression (eSVs; variants with at least one expression hit at LD ≥ 0.8 in GTEx or Geuvadis, or lead variants).

Next, we used the real and imputed genotypes obtained in the previous sections to investigate the contribution of inversion-like variants to gene expression variation by performing a joint *cis*-QTL mapping analysis using RNA seq data of 45 tissues and 2 cell lines of 67-576 individuals from GTEx (GTEx Consortium 2020) and lymphoblastoid cell lines (LCLs) of 358 EUR and 89 AFR individuals from the Geuvadis project (Lappalainen et al. 2013). Specifically, we searched for associations between the expression levels of genes with their transcription start site (TSS) within a ±1-Mb window from our inversion-like variants and all other variants identified in the same region in the corresponding dataset (including mainly SNPs and small indels), considering only common variants (MAF ≥ 0.01 in the available samples). Except for the PCR and BreakSeq genotypes of Geuvadis samples, the analysis was largely based on the imputed data described before. In addition, for 14-18 NAHR inversions that cannot be imputed accurately, a reduced analysis in a smaller number of Geuvadis samples with PCR genotypes was done. Variants in our dataset were strictly identified as eQTLs only when they show significant associations with expression levels of a gene in a particular tissue and they were the lead variant or in high LD (*r^2^* ≥ 0.8) with lead markers (Table S7).

In the GTEx data, we identified a total of 23,935 eQTL associations across all tissues, which correspond to 12,867 variants of different types associated with the expression of 3,348 genes close to inversions or InvDups. Of the 146 inversion-related variants with MAF ≥ 0.01 that could be analyzed in this dataset (30 NAHR inversions, 67 NH inversions, 47 InvDups, and 2 deletions), we found 416 associations with gene expression levels in a given tissue with 26 different variants that were considered to be lead, which included cases almost completely linked with the top variant (*r^2^* ≥ 0.95) to take into account the effect of possible variation or imputation errors (Table S7). This represents basically one fifth of the inversions assessed in Europeans and it highlights the substantial potential consequences that inversions can have in the genome. Interestingly, our analysis revealed that a higher proportion of NAHR inversions (50.0%, 15/30) was identified as eQTL compared to NH inversions (7.5%, 5/67) and InvDups (12.8%, 6/47) (Figure 6A). In addition, the three types of variants were more likely to be involved in gene expression changes than SNPs in the same regions, with enrichment odds ratio (OR) ranging between X and Y, even considering just those showing the highest association to avoid any bias towards inversions and InvDups (Figure 6B). Moreover, when the observed associations were classified by gene category, apparently NAHR inversions were more frequently associated with expression changes in pseudogenes (Figure 6C). Regarding the gene expression analysis in LCLs from the Geuvadis cohort, we found nine associations with three NAHR inversions (17q21.31, HsInv0124 and HsInv1110) acting as lead variants in EUR, whereas there was only one involving the gene *RP11-326C3* and HsInv0124 in the much smaller AFR sample (Figure 6A; Table S10). Conversely, no significant associations were found in the limited analysis of non-imputable NAHR inversions using only PCR genotypes. Therefore, all the rest of the results were just based on the GTEx data in multiple tissues.

**Figure 6.**
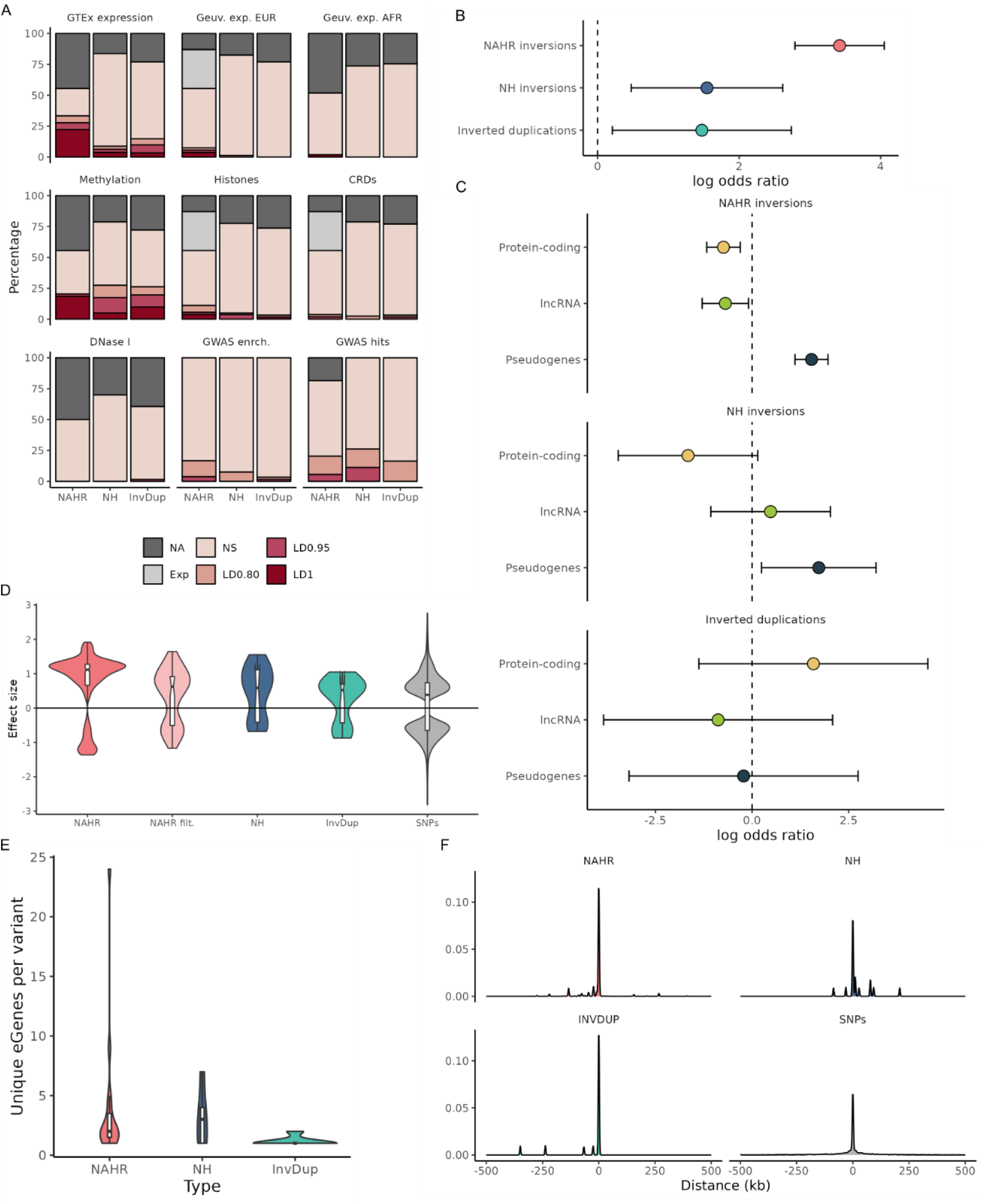
Association of inversion-like variants with gene expression levels and other types of functional signatures. **A.** Proportion of the whole set of NAHR and NH inversions (NAHR/NH Inv) and inverted duplications (InvDups) that show significant results in different functional analyses, representing the strength of the linkage disequilibrium (LD) with the potential lead variant in different shades of red (*r*² = 0.8-0.95, *r*² = 0.95-1, or *r*² = 1). Variants not analyzed due to low frequency or low imputability are indicated as NA and those with no significant results as NS. For data from 1KGP samples, some non-imputable NAHR inversions could be analyzed using just the small fraction of available experimental genotypes (Exp), although no significant associations were found. **B.** Enrichment of inversion-like variants identified as potentially lead eQTLs in GTEx data (LD with top hits of *r*² ≥ 0.95), represented by the Log odds ratio (colored dots) with error bars indicating the 95% confidence intervals. **C.** Enrichment of expression effects of inversion-like variants on different types of genes in GTEx data (LD with top hits of *r*² ≥ 0.95), represented by the Log odds ratio (colored dots) with error bars indicating the 95% confidence intervals. **D.** Distribution of GTEx gene expression effect sizes of different types of variants, with effects of NAHR inversions 17q21.31 and HsInv1110 excluded in NAHR filt. **E.** Distribution of the number of different genes with expression associated to each type of inversion-like variant (eGenes) across GTEx tissues. **F.** Distribution of the distances from the different types of variants identified as eQTLs in this study to their affected genes. For boxplots, the central line represents the median, the box the inter-quartile range (IQR) including the second and third quartiles, and the whiskers extend to the minimum and maximum data values within 1.5 times the IQR.

Given their relatively large size, inversion-like variants could be more likely to influence the expression of multiple nearby genes compared to SNPs. In fact, the mean number of genes affected by our variants was 4.1 for NAHR inversions, 3.2 for NH inversions, and 1.1 for both InvDups and SNPs, showing significant differences between NAHR inversions and InvDups (Mann-Whitney U test: *P* = 2.29 x 10^-2^) (Figure 6E). Our estimates were also higher than the average of 1.8 affected genes per SV reported by (Scott et al. 2021). The elevated number of genes associated with NAHR inversions was driven in part by two of the largest inversions, 17q21.31 (589.2 kb) and HsInv1110 (76.5 kb), which accounted for 61.6% and 13.9% of all inversion-associated results and were linked to expression changes of 24 and 9 genes, respectively. When they were excluded, the mean number of unique genes altered by NAHR inversions lowered to 2.2, but still differed significantly from that of InvDups (Mann-Whitney U test: *P* = 0.040). Consistent with this, inversions and InvDups associated with expression changes were larger in size than those that are not (185.0 vs 16.5 kb; Mann-Whitney U test: *P* = 3.20 x 10^-3^). Thus, these results suggest that longer inversions, which coincide with those generated by NAHR, tend to have more gene expression effects than shorter ones.

Regarding the magnitude of the effects, their distribution was similar across the different variant types, although inversions tend to have bigger effect sizes that were biased towards positive values, particularly for NAHR inversions (Kolmogorov-Smirnov test: NAHR vs NH *P* = 1.12 x 10^-2^; NAHR vs InvDups, *P* = 3.24 x 10^-7^; NAHR vs SNPs, *P* < 2.2 x 10^-16^) (Figure 6D). Remarkably, when the 17q21.31 and HsInv1110 inversions were excluded, the distribution became more symmetrical (Figure 6D), differing only from that of SNPs (Kolmogorov-Smirnov test: NAHR vs SNPs, *P* = 3.76 x 10^-3^). This suggests that the bias towards increased gene expression in NAHR inversions would be primarily driven by these two inversions. The global upregulating effects of the 17q21.31 inversion on nearby genes were attributed in part to the duplications existing in the H2 haplotype and involved a high number of pseudogenes (Campoy et al. 2022). Similarly, In the case of HsInv1110, most of its effects were observed in pseudogenes, which are probably subject to less stringent regulation, However, the exact regulatory mechanisms driving these effects are not clear.

Actually, a high proportion of eQTLs identified in our analysis are intronic or intergenic, including 91% of the SNPs, 60% (3/5) of the NH inversions, and all the InvDups, whereas 86.7% (13/15) of the NAHR inversions overlap with exons, as expected by their larger size (Figure 5C). These findings align with prior studies, which reported that 82.3% of unique SV-eQTLs did not intersect with any exons of the affected gene (Scott et al. 2021). When the distances between the eQTL variants and their genes are calculated, a greater average distance was found for SNPs with respect to all types of inversion-related variants (NAHR inversions: 26.2 kb; NH inversions: 34.7 kb; InvDups: 35.7 kb) (Figure 6F). This is consistent with inversions, especially those generated by NAHR, containing in many cases genes (corresponding to a distance to the TSS of zero).

We also examined the number of tissues in which eQTLs for a given gene were identified. A total of 67 eQTLs associated with inversion-related variants (Inv-eQTLs) were found in fewer than five tissues. Notably, Inv-eQTLs present in 10 or more tissues (11) are exclusively mediated by three of the largest NAHR inversions: the 17q21.31 inversion, HsInv1117 and HsInv1110. However, as already mentioned, the majority of these widespread associations involve pseudogenes (8/11). On average, expression changes associated with inversion-related variants were observed in 3.6 tissues for protein-coding genes, 3.1 for lncRNA genes and 7.9 for pseudogenes. This suggests that expression changes in protein-coding genes, which are expected to have more serious functional consequences, might be subjected to a tighter tissue-specific regulation. In contrast, pseudogene expression changes, less constrained by natural selection, may persist across tissues without being eliminated due to negative consequences.

As a last step, we tried to estimate how many of the reported inversion-related gene expression effects were novel or had been identified in previous GTEx analyses (GTEx Consortium 2020). Due to presence of multiple SNPs in high LD with the inversion and InvDup alleles, some already-known GTEx eQTLs may in fact be driven by SVs not included in those analyses. In contrast, the effects of recurrent inversions in low LD with neighboring variants have been probably missed, unless they were accurately imputed in the dataset. To address this, we explored the 742 associations in which inversion-like variants were lead or in strong LD (*r*² ≥ 0.8) with the corresponding lead variant. In 92 out of the 742 associations (12.4%) we did not find a matching eQTL within the GTEx catalog involving the same gene and tissue as those identified in our analysis (Table S9). In total, we found potentially new regulatory associations for 26 genes mediated by 12 different inversions (8 NAHR, 4 NH) in different tissues, the majority of which belong to the 17q21.31 and HsInv1110 inversions (65/92, 70.7%). For the remaining 650 associations, we determined whether the effect sizes of inversion-like variants were larger than those reported for SNPs. As shown in Figure S2, the proportion of gene expression associations in which inversion-like variants show a larger effect increased with their probability of being the lead variant (measured as the LD between them), Indicating that the potentially greater influence of inversion-like variants in those cases is missed in the available GTEX data. Therefore, this emphasizes the importance of the careful analysis of previously unknown variants to characterize better the determinants of gene expression variation.

Nevertheless, it is important to take into account that, in general, three main factors influenced the discoveries in our analysis (Figure 6A): (1) the number of samples in the dataset; (2) the number of tissues with expression data; and (3) inversion genotype reliability. First, larger cohorts of samples provide higher power to detect significant differences and they allow the inclusion of low-frequency variants that otherwise would be excluded from the analysis, resulting in more inversion-associated changes. This is illustrated by the highly significant correlation observed between the number of eQTL associations and sample size across all GTEx tissues (Figure S3). Also, diverse ancestries are crucial to identifying the effects of population-specific SVs. Second, the analysis of a wide set of tissues allows to interrogate more genes with tissue-restricted expression and identify tissue-specific regulatory effects. For example, many less gene expression associations were observed in Geuvadis LCLs compared to GTEx despite the large sample size (Figure 6A). Finally, due to the difficulty of imputing reliably recurrent NAHR inversions, a significant proportion of them were not included in the gene expression analysis or could be analyzed just in a reduced number of samples (Figure 6A), so their functional consequences remain unexplored.

### Effects on epigenetic modifications and chromatin organization

Besides gene expression levels, we also examined the effect of inversion-related variants on other relevant genomic functional signatures involved in gene regulation, such as DNA methylation, histone modifications and chromatin accessibility data available in a subset of GTEx tissues and 1KGP LCL samples (see Methods). As before, the analysis was based on using the available genotypes to search for cis-QTLs close to inversion regions for each type of molecular phenotype and identify those that are lead variants or strongly linked (*r*^2^ ≥ 0.95) with the lead variants.

First, when we tested the associations with DNA methylation profiling of more than 750,000 genomic positions (CpGs) in several of the GTEx tissues (Oliva et al. 2023), we found 29,588 significant associations, 0.9% of which (258/29,588) involved potentially lead inversions or InvDups (Table S11). In total, 27.7% (38/137) of the tested inversion-like variants were associated with methylation levels, corresponding to 11 NAHR inversions (36.7%), 14 NH inversions (22.2%) and 13 InvDups (29.5%), with similar proportions among the different types of variants (Figure 6A). However, as for the eQTLs, more NAHR inversions showed an effect in methylation, and in almost all cases the inversion was lead or completely linked with the lead variant (*r*^2^ = 1). Therefore, these results provide insights into inversions that may influence gene expression through epigenetic modification of regulatory elements.

Second, we analyzed the influence of our SVs on three different histone modification marks (H3K27ac, H3K4me1 and H3K4me3) obtained from 1KGP LCLs. Of the 1,974 reported significant associations, in 13 (0.7%) the potentially lead variants correspond to inversions (3 NH and 3 NAHR) or InvDups (1) (Figure 6A; Table S11). Interestingly, this included four associations with inversion HsInv0124 (3) and HsInv0030 (1) and one with InvDup HsInv1454 in which they acted directly as lead markers. With respect to the type of histone mark, we found seven associations with changes in H3K4me1 (typical of enhancers), four with H3K27ac (usually marking active enhancers and promoters) and two with H3K4me3 (enriched at the promoters of active genes) (Table S11). Histone modifications have also been used to identify cis-regulatory domains (CRDs) that constitute finer organization genomic regions in which genomic elements coordinately regulate gene expression (Delaneau et al. 2019). Consistent with the observed changes in histone modifications, we found changes in CRD activity in LCLs associated with inversion HsInv0124 and InvDup HsInv1454 as lead variants (Figure 6A).

Finally, we tested whether inversion-like variants are linked to changes in chromatin accessibility using DNase I hypersensitivity sequencing data from 54 African LCL 1KGP samples from the (Degner et al. 2012). Only one significant association with InvDup HsInv0997 linked with the lead marker (*r*² ≥ 0.95) was found (Figure 6A), probably in part due to the small sample size.

### Inversions and phenotypic traits

To explore the contribution of inversion-like variants to phenotypic variation, we exploited the known GWAS hits collected in the NHGRI-EBI GWAS Catalog (MacArthur et al. 2017) using two complementary approaches. First, we investigated if there was an enrichment in the number of trait-associated signals in the inversion and flanking regions (± 20 kb) compared to what should be expected by chance. Our analysis revealed a 2.20-fold enrichment of GWAS hits within inversion regions compared to the rest of the genome (*P* = 0.02), which increased to 3.43-fold for inversions exceeding 100 kb (Figure 7A). The enrichment is mainly driven by 15 variants, including 7 NAHR inversions, 6 NH inversions, and 2 inverted duplications (Figure 3; Figure 7B). In particular, the 8p23.1 and 17q21.31 inversions, which have been extensively characterized and are associated with multiple phenotypic effects (Campoy et al. 2022), showed the highest GWAS signal enrichment. When considering the mechanism of origin, a 3.08-fold enrichment was observed exclusively for NAHR inversions (*P* = 0.01), consistent with their larger size and the results of these two inversions, whereas NH Inversions and InvDups overall showed a distribution of GWAS hits similar to the rest of the genome (Figure 7A). Some of these GWAS signals can be attributed to the presence of the inversion and for example those close to HsInv0124, which is located at the *IFITM* locus involved in innate immunity, are related to distinct blood cell counts, bone mineral density for HsInv0095, mental abilities and psychological conditions (intelligence, attention deficit hyperactivity disorder, educational attainment or adventurousness) for HsInv0174, and IgG glycosylation patterns for HsInv1321.

**Figure 7.**
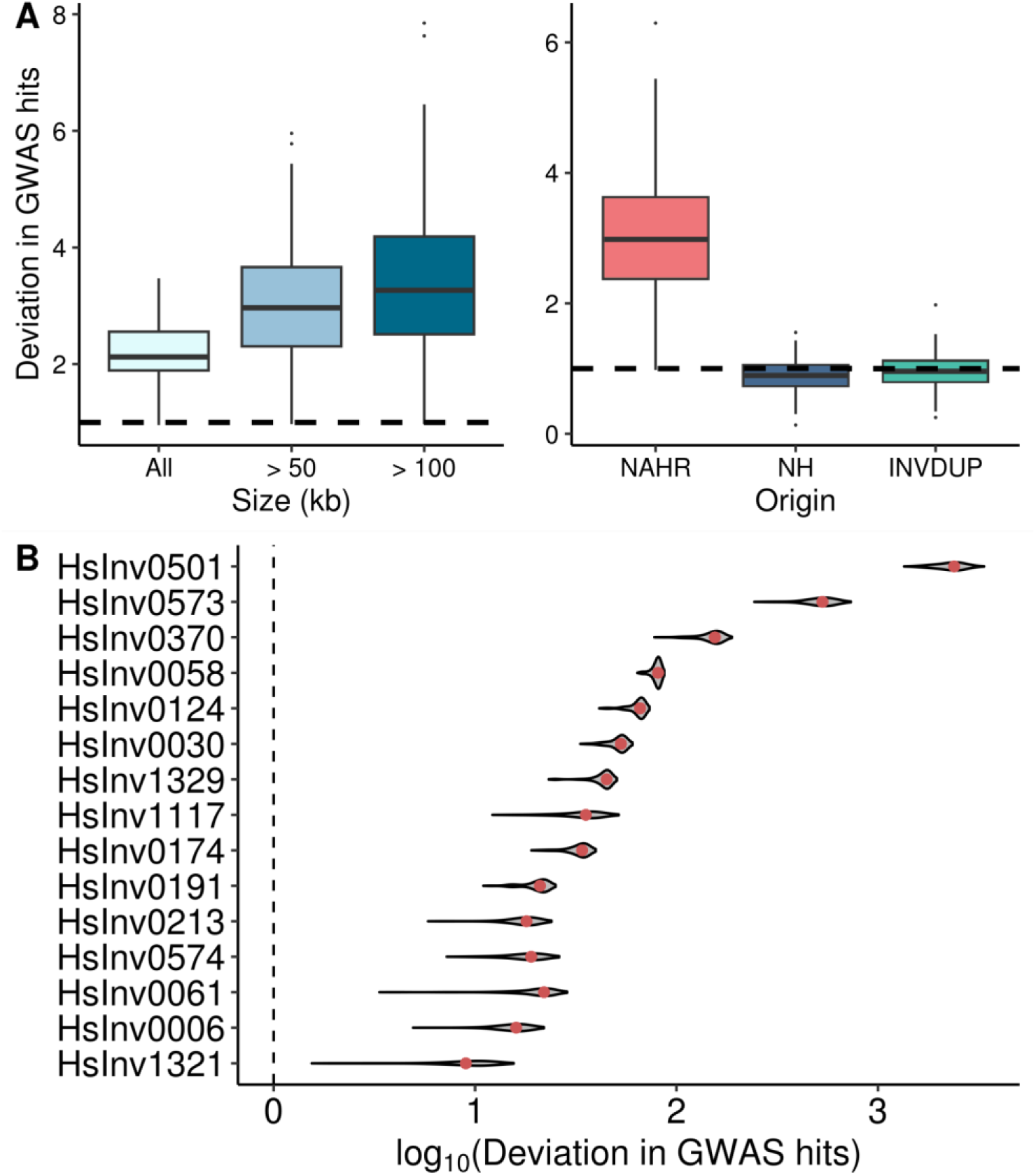
Enrichment of GWAS Catalog trait-associated signals on inversion-like variant regions. **A.** Fold enrichment of observed GWAS hits within inversion-related variants and flanking regions (± 20 kb), compared to those expected in random genomic regions of the same size, across different groups of inversions classified by size (left; All, n = 193; > 50 kb, n = 26; > 100 kb, n = 15) or type of variant. **B.** Summary of the distribution of the differences in the number of observed minus expected GWAS signals (base-10 logarithm transformed) for the 15 individual inversions and InvDups showing a significant GWAS hit enrichment (nominal *P* < 0.05, one-tailed permutation test), represented as the 95% one-side confidence interval with the median indicated by a red dot.

Second, by searching for SNPs in high LD (*r*² ≥ 0.8) with the inversions and InvDups that show significant GWAS signals in the GWAS catalog, it was possible to identify possible phenotypic effects of inversion-like variants. In total, 42 variants were associated with GWAS signals, with similar proportions for NAHR (11/33, 33.3%) or NH (21/80, 26.3%) inversions and slightly less for InvDups (10/61, 16.4%) (Figure 6A). This represents a considerable enrichment of GWAS associations, especially in the case of recurrent NAHR inversions for which LD with SNPs tend to be low and they can be interrogated just in certain populations. However, although the 17q21.31 inversion alone accounted for 487 GWAS hits, the number of associated traits did not differ significantly across inversion types (One-tailed Kolmogorov-Smirnov test, *P* = 0.300). Apart from the well-studied 8p23.1 and 17q21.31 inversions (Campoy et al. 2022), for which the updated analysis identified numerous new associations, including diverse lipid-related traits for the 8p23.1 inversion that could be linked to its impact on obesity, HsInv0786 stands out as the variant with the highest number of associations (37), followed by HsInv1075 (23), HsInv0191 (17), HsInv1117 (15) and HsInv0174 (15) (Figure S4, Table S12). Interestingly, in some cases the associated traits show a clear biological connection, like those of HsInv0191 that are all linked to cardiovascular health. In addition, we have also found new interesting GWAS associations, like that of HsInv0379, that disrupts the *ZNF257* gene and creates a fusion transcript, with diabetes in Asian individuals (Puig et al. 2015b; Spracklen et al. 2020). Thus, these results support that inversions could have a relevant role on phenotypic traits.

### Integrative analysis of inversion functional effects

Lastly, we have performed an integrative analysis of all the available data, which is crucial to dissect in detail the consequences of some of the variants at the molecular level and contribute to a better understanding of the genetic basis of the associated traits. Interestingly, 56.2% of inversion-like variants showed at least one significant result in the different analyses (*F_ST_*, effects on genes, gene expression, DNA methylation, histone modifications and CRDs, or GWAS signals) (Figure 3), emphasizing their potentially important genomic impact. In addition, the distribution of these functional signals was not random, with effects accumulating in certain variants, including 13 that showed significant results in four or more analyses. Consistent with their larger size, NAHR inversions tended to have a higher number of effects and an enrichment of variants with many effects (≥4) (14.9%, 7/47: HsInv0030, HsInv1117, HsInv0124, HsInv0573, HsInv0786, HsInv1110 and HsInv0348) compared to NH inversions (4.5%, 3/67: HsInv0191, HsInv0058 and HsInv1065) and InvDups (6.4%, 3/47: HsInv0997, HsInv0310 and HsInv1454) (Figure 3), which is especially remarkable considering that for many NAHR inversions the power of some analysis is limited. These 13 variants were all associated with gene expression and DNA methylation differences, with 10 of them showing also GWAS hits association or enrichment, 10 involving changes in histone modifications and 6 in CRDs, 5 having high *F_ST_* values among populations, and 4 affecting gene exons (Figure 3).

This analysis confirmed the pervasive functional role of some already characterized inversions, extending their possible effects in gene expression and phenotypic traits, and identifying novel associations with epigenetic changes. That is the case of the well-known 17q21.31 inversion and HsInv0786 (Campoy et al. 2022; González et al. 2014; Puig et al. 2020; González et al. 2020). Similarly, it was previously reported that HsInv0124 was the lead eQTL of several long non-coding RNAs (lncRNAs) overlapping the *IFITM* genes and it showed significant frequency differences between populations (Giner-Delgado et al. 2019). Here we validated these results and found additional associations with DNA methylation, histone modifications and changes in CRDs (Figure 3), which may play a role in the possible regulatory mechanism of the inversion (Lerga-Jaso 2019). A different case is that of HsInv0058, located within one of the functionally diverse Major Histocompatibility Complex (MHC) haplotypes on Chr. 6 accumulating remarkable sequence and structural differences (Giner-Delgado et al. 2019), which are probably responsible for the many effects linked to the inversion (Figure 3). Moreover, our analysis replicated and identified new effects of other inversion candidates highlighted before, such as HsInv0006 and HsInv0201 (Giner-Delgado et al. 2019). For example, in HsInv0006, which showed possible signals of positive selection in AFR (Giner-Delgado et al. 2019), we found associations with red cell distribution width, meat related diet and several epigenetic changes that could be related to variation of expression levels of many genes in this region in different tissues, although the inversion is not the lead eQTL (Figure 3). Also, in the old HsInv0201 inversion that could be maintained by balancing selection (Giner-Delgado et al. 2019), the GTEX results indicate that the inversion acts as lead variant of the expression of two additional neighboring genes and methylation of a flanking CpG position (Figure 3; Table S9; Table S11).

One known inversion with potentially important consequences that we have examined in more detail is HsInv0030, which exchanges the promoter and first exon of the *CTRB1* and *CTRB2* genes in the ancestral *O2* orientation (named as *CTRB1-Q* and *CTRB2-Q*) generating new chimeric gene combinations in the derived *O1* reference orientation (named as *CTRB1-R* and *CTRB2-R*) (Figure 8A), and has been linked to expression and activity differences of the encoded chymotrypsinogens in pancreas that could be involved with its association with non-alcoholic pancreatitis susceptibility (Rosendahl et al. 2018; Giner-Delgado et al. 2019). Consistent with these results, we have found that the inversion is the lead variant of *CTRB1* upregulation and *CTRB2* downregulation in pancreas (Figure 8B; Table S9), together with different epigenetic changes in CpG methylation and histone modifications in several other tissues, although unfortunately information from pancreas was not available (Table S11). By mapping pancreas RNA-seq reads from seven *O1/O1* and *O2/O2* homozygote individuals to the specific nucleotide positions in the first and last exons of the genes that allow us to differentiate the diverse *CTRB* transcripts, we found that the observed expression changes are likely related to the exchange of promoters, but unlike *CTRB2-Q* and *CTRB1-R*, *CTRB2- R* shows a very strong expression increase compared to *CTRB1-Q* despite sharing the same promoter (Figure 8C), suggesting the existence of additional regulatory processes.

**Figure 8.**
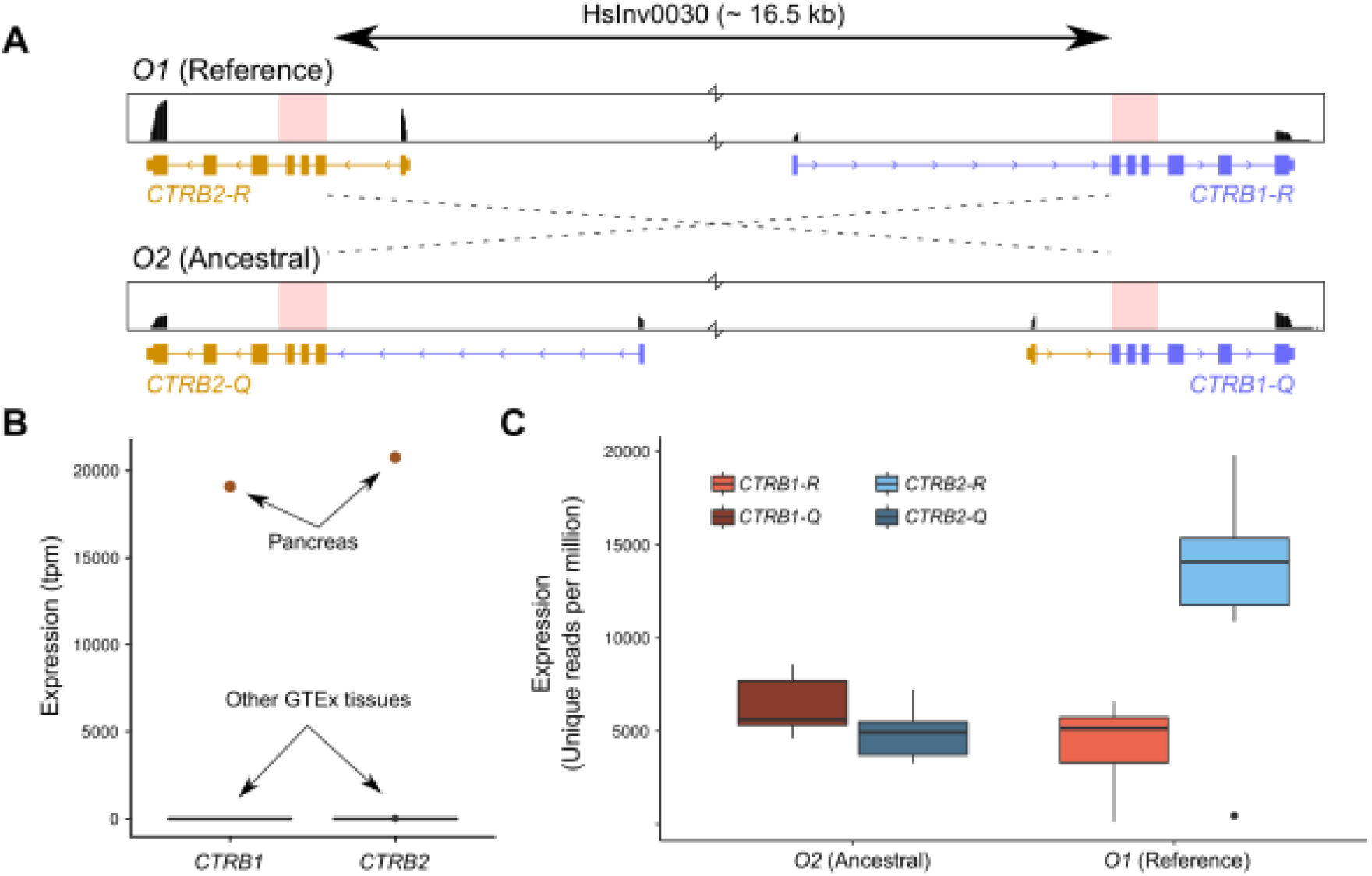
Molecular consequences of inversion HsInv0030. **A.** Gene structure and RNA-seq profiles from pancreas GTEx reads for seven *O1/O1* or *O2/O2* individuals mapped to the first and last exons of *CTRB1* and *CTRB2* genes that are exchanged by the inversion (indicated by dashed lines) mediated by NAHR between the highly identical SDs (marked in pink). **B.** Average expression levels of *CTRB1* and *CTRB2* genes in different tissues, showing that they are exclusively expressed in pancreas. **C.** Detailed analysis of expression changes of the different *CTRB* transcripts associated with the HsInv0030 inversion. Box plots represent the expression levels for the seven individuals analyzed in **A**.

Nevertheless, among the inversion-like variants displaying four or more different associations there are also several interesting novel candidates. This includes three InvDups, although they are relatively distant from known gene exons and, except for for HsInv1454, mostly they are not lead variants of the observed differences (Figure 3). Contrarily, the new candidate inversions tend to show a clearer link to their possible functional effects at the molecular level. For example, HsInv1065 is a complex rearrangement comprising two 1.8 and 2.9-kb inversions, a small duplication and four relatively large deletions affecting four olfactory receptor genes, which could explain some of the epigenetic changes for which it was the lead variant (Figure 3). Another intriguing inversion due to its association with different cardiovascular traits and heart disease was HsInv0191, located within an intron of the gene *SYNPO2L,* that was a lead variant of *ZSWIM8* gene expression and DNA methylation of two CpG sites in non-heart tissues (Figure 3; Table S9; Table S11). Actually, the inversion is in high LD with a group of tightly linked SNPs that have been shown to modulate the differential usage of *SYNPO2L* isoforms and the expression of *SYNPO2L* and *MYOZ1* in opposite directions, which together encode proteins essential for the maintenance and stabilization of muscle fibers, including cardiac muscle, and are probably responsible of the GWAS hits (Wass et al. 2024).

However, one of the best new examples of inversions causing clear molecular changes and being associated with phenotypic traits is HsInv1117, a low frequency inversion that has risen in frequency from 1.1-2.8% in AFR and EAS, to 12.9-15.4% in EUR and SAS, although population differences are not significant (Figure 3; Table S4; Table S5). The inversion was apparently generated by a unique NAHR event between two 94.8% identical SDs of 47.5-57.1 kb that contain the *AKR1C1* and *AKR1C2* genes, switching their position and exchanging the 5’ upstream region flanking the TSS of the main isoforms (Figure 9A). Our analysis showed that the inversion upregulates *AKR1C2* mRNA levels in practically all tissues, and it was the lead variant of expression changes of a few other genes in the region, including a more modest decrease of *AKR1C1*, and methylation levels in one CpG in specific tissues (Table S9; Table S11). Consistent with these results, by mapping LCL RNA-seq reads, we found that in the *O2* orientation there was a considerable expression increase of a long transcript containing six additional upstream exons plus the rest of the *AKR1C2* gene (Figure 9A), which was now the highest expressed isoform. Similar analysis in two other tissues showed that in adipose tissue, only the shorter isoform of both genes was expressed at equivalent levels in the three inversion genotypes. Conversely, in testis, the expression of the longer isoform resulted in a decrease of *AKR1C1* transcript levels in the *O2* orientation together with a much higher *AKR1C2* increase, as was observed in LCLs (Figure 9B). This illustrates that HsInv1117 acts through diverse gene regulation mechanisms, which are tissue specific. In addition, by LD with SNPs, the inversion was associated with a decrease of the waist-to-hip ratio and height and an increase in *AKR1C1* protein levels (Figure S4; Table S12). Interestingly, the encoded AKR1C1 and AKR1C2 aldo-keto reductases have different functions in metabolism of steroid hormones, which are important drivers of body shape.

**Figure 9.**
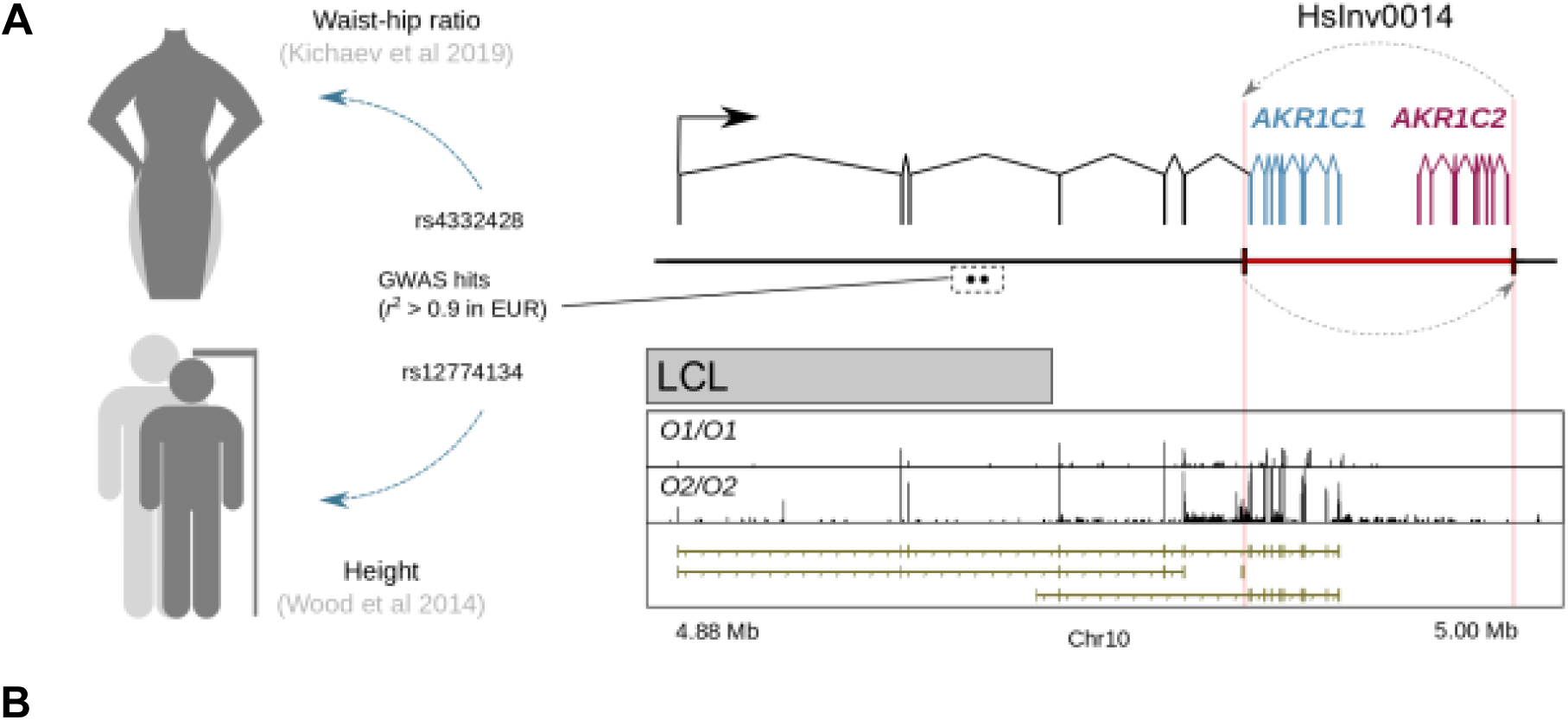
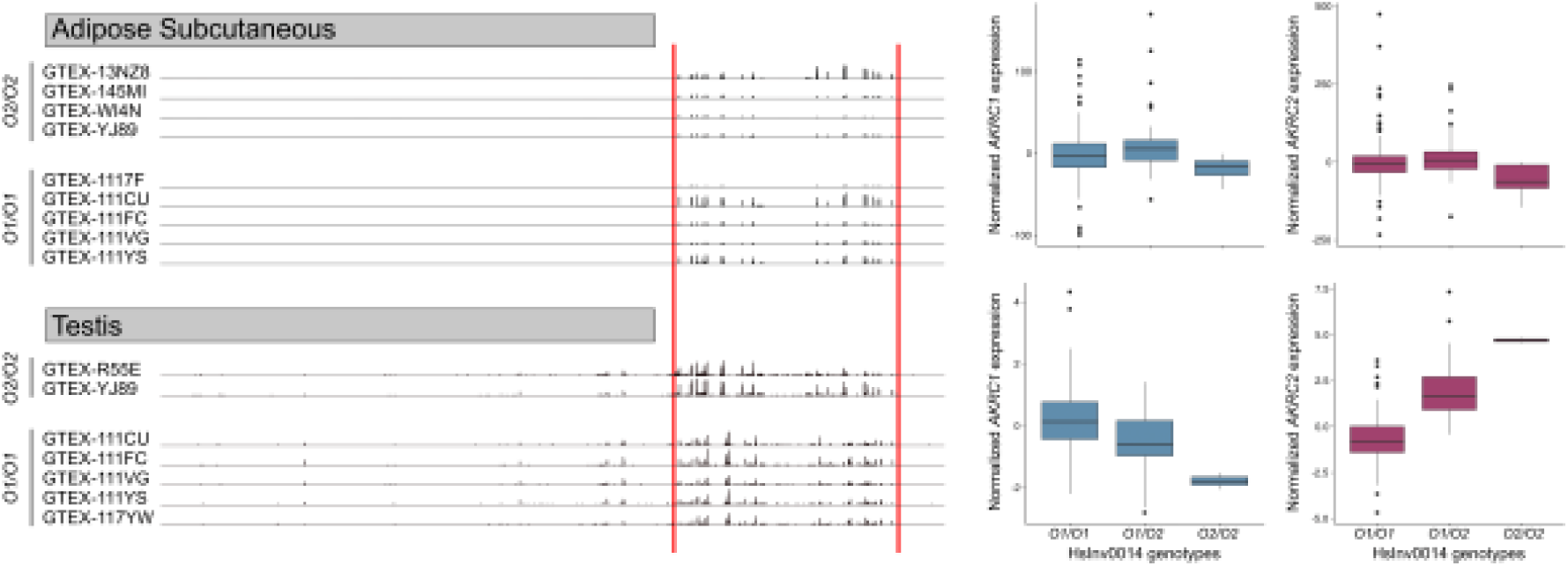
Summary of HsInv1117 inversion effects on *AKR1C1* and *AKR1C2* expression and its association with body shape. **A.** Remodeling of gene *AKR1C1* (blue) and *AKR1C2* (red) by the inversion (dashed arrow), showing the extended isoform (black) upregulated in O2 chromosomes and the RNA-seq profiles from LCL reads for *O1* and *O2* homozygotes. HsInv1117 is in high LD in Europeans (EUR) with GWAS variants associated with waist-hip ratio and height that are located within an intron of the long isoform. **B.** Tissue-dependent regulation of *AKR1C1* and *AKR1C2* expression by the HsInv1117 inversion. RNA-seq read profiles from adipose subcutaneous tissue and testis of several *O1* and *O2* homozygotes in the HsInv1117 region (left), and boxplots of resulting *AKR1C1* and *AKR1C2* expression levels by inversion genotype (right), showing diverse effects of HsInv1117 on gene regulation depending on the tissue assayed. In both panels, inversion breakpoints are indicated by red lines.

Moreover, HsInv1110 was another novel strong candidate inversion that is relatively large, accumulates a high number of different effects, and is only present at low frequency in EUR (Figure 3). In fact, the inversion acted as lead eQTL of 57 expression changes across 9 genes in 26 different tissues, including *ENSG00000224905/AP001347.6* (*LIPI-AS*), a lncRNA antisense to the protein-coding gene *LIPI* that is disrupted by the inversion (Figure 10A; Table S9). Also, consistent expression associations were found in additional tissues for all genes except one when the LD threshold with the lead variant was lowered to *r*^2^ ≥ 0.8. Besides a decrease in expression of *LIPI* and *LIPI-AS* in several tissues, the expression changes comprised upregulation of seven pseudogenes, which are mostly located in the upstream region of *LIPI-AS*. Interestingly, unlike the 17q21.31 and HsInv0786 inversions, which maintain relatively-divergent haplotypes of highly linked variants (Puig et al. 2020; Campoy et al. 2022), HsInv1110 does not show high LD with other variants, and tends to be the sole top variant or its association *P* value is almost indistinguishable of that of the top SNP, suggesting that it is likely responsible of the observed changes (Figure 10B). One possibility is that the reorientation of the *LIPI-AS* promoter and first exon may influence the observed overexpression of pseudogenes proximal to the first breakpoint. In addition, the *LIPI-AS* antisense RNA may play a role in regulating *LIPI* expression and/or translation, a process potentially disrupted by the inversion, and both transcripts showed a positive correlation of expression levels in different tissues, with the decreased expression of *LIPI-AS1* in the *O2* orientation being associated to lower *LIPI* expression levels (Figure 10C). Finally, the inversion was also associated with DNA methlylation of multiple CpGs throughout the region, which mostly showed decreased methylation levels, and it is highly linked (*r*² = 0.89) to the lead variant associated to elevated H3K4me3 levels near the TSS of the *ANKRD20A18P* pseudogene, providing a possible mechanism for its increased expression (Figure 10A; Table S11). Interestingly, LIPI encodes a phospholipase that hydrolyzes phosphatidic acid into lysophosphatidic acid and has been associated to familial hypertrigliceridemia susceptibility (Wen et al. 2003). However, since there are no linked SNPs with *r*² ≥ 0.8 in EUR, it has not been possible to associate the inversion to any GWAS hits so far.

**Figure 10.**
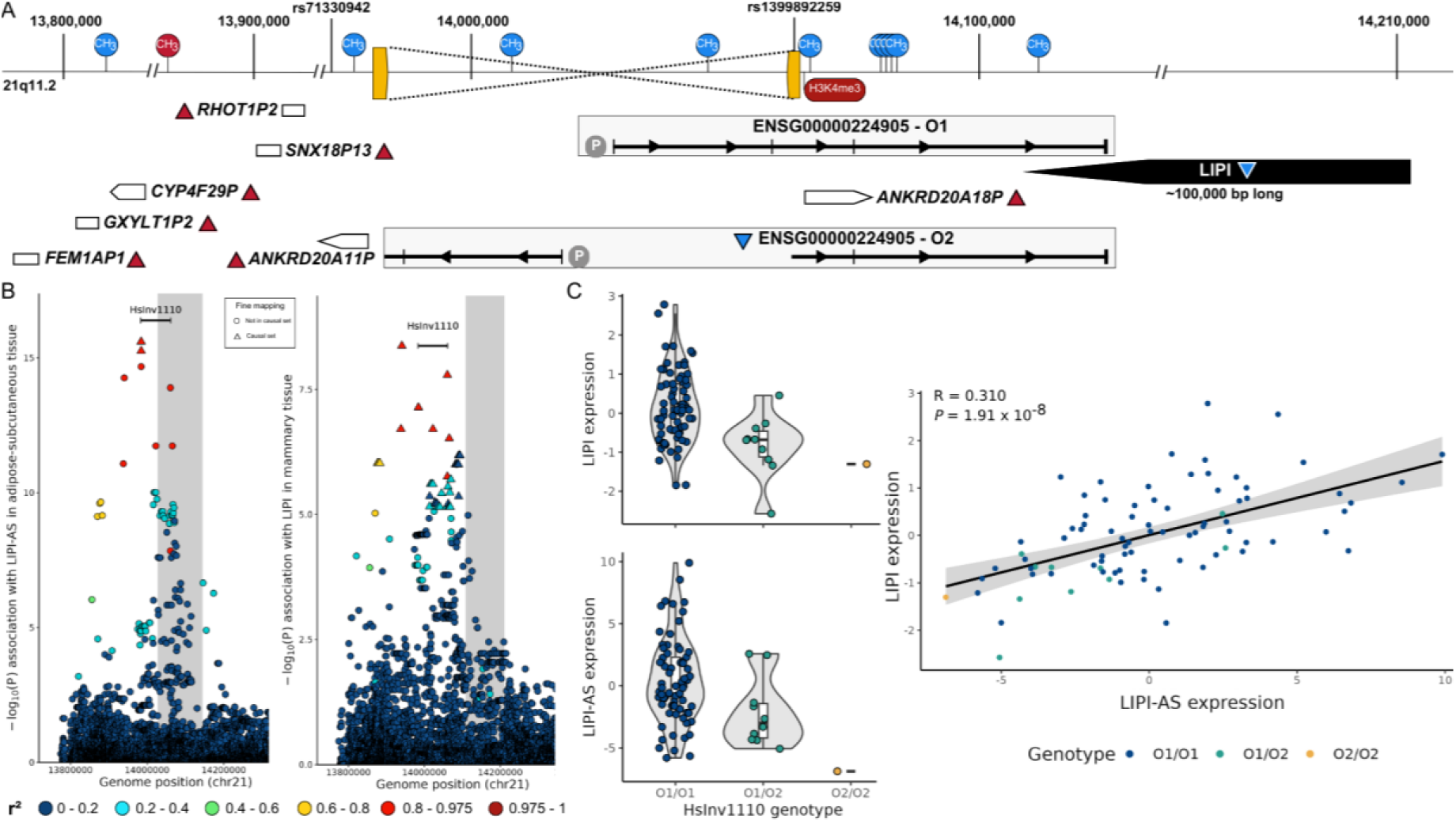
Summary of HsInv1110 inversion effects on gene expression and DNA epigenetic changes. **A.** Diagram of the Chr. 21 genomic region harboring the HsInv1110 inversion originated by recombination between inverted SDs (yellow blocks), which in the *O2* orientation disrupts the ENSG00000224905 (*LIPI-AS1*) long non-coding RNA gene (represented within the gray rectangle with exons as vertical lines and the possible promoter as P). The other genes in the region are represented as white (pseudogenes) or black (*LIPI* protein coding gene) rectangles, with upward (red, increase) or downward (blue, decrease) triangles indicating changes in expression levels in the *O2* orientation. Arrows within the genes indicate the direction of transcription. CpG methylated sites (CH3) and a histone modification peak (H3K4me3) showing higher (red) or lower (blue) levels in the *O2* haplotypes are shown above and below the diagram, respectively. **B.** Manhattan plot of logarithm-transformed GTEx eQTL *P*-values of the HsInv1110 inversion (represented by a black bar) and surrounding SNPs (circles and triangles colored according to their LD with the inversion) for *LIPI-AS1* (left) and *LIPI* (right) (with gene position indicated by the gray highlight) in adipose-subcutaneous and breast tissue. **C.** Boxplots of rank normalized *LIPI* and *LIPI-AS1* expression according to HsInv1110 inversion genotype (indicated by circles of different colors) (right) and correlation between the expression of both genes (left), with colored circles indicating the HsInv1110 inversion genotype, with Spearman correlation (*R*) and its *P* value indicated within the graph. The decreased expression of *LIPI-AS1* in the *O2* haplotype could explain the change in *LIPI* expression in the same haplotype (R = 0.310; *P* value = 1.91 x 10^-8^).

Therefore, our detailed analysis based on accurate genotypes and reliable imputation of inversion-like variants, has allowed us to show that a high proportion of this type of variants have important functional effects, studying many of them for the first time. In addition, together with some of the previously discovered inversions, such as HsInv0379 or HsInv1051 (Puig et al. 2015b; Giner-Delgado et al. 2019), it has identified many interesting candidates linking potential effects at the molecular level and associations with significant phenotypic traits, which contribute to expand our current knowledge of the genetic basis of phenotypic characteristics.

## Materials and methods

### Generation of the inversion-like variant dataset

To extend our prior catalogue of human inversions (Martínez-Fundichely et al. 2014; Aguado et al. 2014; Vicente-Salvador et al. 2017; Giner-Delgado et al. 2019; Puig et al. 2020) and ensure a good representation of the maximum number of common variants (frequency >1%), we first analyzed a wide diversity of inversions identified using diverse techniques with different precision levels (Korbel et al. 2007; Kidd et al. 2008; Chaisson et al. 2015; Sudmant et al. 2015; Hehir-Kwa et al. 2016; Huddleston et al. 2017), available at the InvFEST database when the project started. As part of InvFEST analysis, similar predictions within and between studies were merged automatically in independent inversion candidates. Due to the high false positive rate of inversion detection, we included all the variants identified with long reads (Chaisson et al. 2015; Huddleston et al. 2017) and only those identified with short reads that were supposedly validated in the corresponding study (Korbel et al. 2007; Hehir-Kwa et al. 2016), excluding inversions located entirely within repetitive elements or in very complex regions that have changed between the GRCh36 (hg18), GRCh37 (hg19) and GRCh38 (hg38) genome assemblies and those with large IRs (>25 kb) that cannot be assayed (Table S1). In addition, we made an effort to analyze all inversions with a global frequency of >1% from the 1KGP Phase 3 (Sudmant et al. 2015).

To confirm the presence of an inversion or other SVs in the candidate regions that have not been validated yet, we searched for sequences supporting the alternative orientation (*O2*) using the hg38 human genome assembly (*O1*) as a reference. For that, we carried out a BLAST search (Altschul et al. 1990) of the predicted InvFEST region (plus 5 kb flanking sequence) against the whole database of non-redundant human sequences. Moreover, we did a more automatic BLAST analysis of the hg38 coordinates reported in the corresponding study ±1 kb of flanking sequence at each side against the downloaded genomic sequences of 69 human genome assemblies from 35 different individuals available at the NCBI Datasets Genome database (https://www.ncbi.nlm.nih.gov/datasets/genome/), which allowed us to identify variants with ∼1% frequency (Table S2). Briefly, if the top BLAST hit was contiguous and extended over the inversion length plus 1,800 bp out of the 2 kb of flanking sequence, the assembly was marked as *O1*. For those genome assemblies and additional BLAST hits that did not match perfectly the *O1* sequence, we determined if there was an inversion or any other rearrangement and annotated precisely the breakpoints by manual inspection of pairwise BLAST alignments and UCSC Genome Browser annotations (http://genome.ucsc.edu). Any alternative structural allele including at least one inverted segment of >50 bp that is not present anywhere else in the genome was considered an inversion, even if it contains other insertions (such as InvDups) or deletions (Table S1). Similarly, alternative structural alleles including at least a duplication of a sequence of >50 bp present in another genome location in opposite orientation were considered an InvDup, even if they contain other insertions or deletions. Also, this allowed us to identify microhomologies or IRs formed by different types of repetitive sequences at the SV breakpoints and characterize their most likely mechanism of generation. Finally, for a few specific well-supported inversions in which available sequences were not informative, we used other available sequence reads (Sudmant et al. 2015) or generated our own *O2* sequences by PCR amplification to validate the alternative orientation and annotate more precisely the breakpoints. Therefore, in just a small proportion of low-frequency candidate inversions of the studies with a higher number of samples the alternative orientation could not be resolved, which comprises 20 variants with <2.5% global frequency in the 1KGP (Sudmant et al. 2015) and 17 from Hehir-Kwa et al. (2016) (Table 1). For inversions mediated by highly identical IRs defined by fosmid PEM (Kidd et al. 2008, 2010), only individual sequences supporting the inverted orientation that span the IR and those with >2 uniquely mapping fosmid end sequences were analyzed.

### Experimental validation and genotyping of inversion-like variants

Experimental validation of candidate inversions was done by regular PCR or iPCR in a small panel of samples, including whenever possible some individuals in whom the inverted orientation had been detected. Both breakpoints in the *O1* and *O2* orientations were tested for all >50 bp inversions with sequence support and those with IRs at the breakpoints for which PCR assays could be designed, which was limited mainly by the size of the IRs (<25 kb) and the presence of restriction enzyme target sites at both sides. This also involved testing at least one breakpoint of a small set of the sequence resolved InvDups. Simple and multiplex PCRs and iPCRs of each inversion were performed as previously described (Aguado et al. 2014; Vicente-Salvador et al. 2017), with specific primer combinations flanking either the inversion breakpoints (PCR) or the self-ligation sites of circularized molecules (iPCR) from both orientations. Primer sequences are available upon request.

In general, genotyping of inversions was done by multiplex PCR of one breakpoint followed by standard gel electrophoresis in a diversity panel consisting of 95 unrelated human samples of four 1KGP populations with AFR (32 YRI), EUR (31 CEU) and EAS (16 CHB and 16 JPT) ancestries (Puig et al. 2020) (Table S3). In addition, for those NAHR inversions not showing perfect tag SNPs in the previous samples, extended genotyping was done in 136 additional samples from AFR (18 YRI), EUR (14 CEU and 90 TSI) and EAS (7 CHB and 7 JPT) ancestry (Table S3). Finally, a few low frequency or especially interesting inversions were genotyped in additional samples previously used for inversion characterization (Giner-Delgado et al. 2019). Genomic DNAs from lymphoblastoid cell lines (LCLs) were obtained directly from the Coriell Cell Repository (Camden, NJ, USA) or had already been isolated (Giner-Delgado et al. 2019).

Furthermore, 143 candidate variants (79 NH inversions, 61 InvDups and 3 deletions) without highly-identical repeated sequences at their breakpoints were genotyped bioinformatically in the 3,202 1KGP-HC samples (Byrska-Bishop et al. 2022) using a modified version of the BreakSeq pipeline (Lam et al. 2010; Giner-Delgado et al. 2019; Lucas-Lledó et al. 2014; Vicente-Salvador et al. 2017), which was extensively optimized to identify sequences spanning simple breakpoints from this dataset containing longer and many more reads than the original 1KGP data (Sudmant et al. 2015). Reads of the target region (±1 kb) and unmapped reads were extracted from the 1KGP-HC BAM files and they were mapped with Bowtie2 (Langmead and Salzberg 2012) to a breakpoint library formed by 300-bp probe sequences centered around the breakpoints of the inversion in both orientations or the insertion or deletion positions with and without the extra sequences for InvDups and other SVs (including between two and eight probe sequences for each variant depending on its complexity and the breakpoint characteristics). To ensure that results were specific, reads were required to match at least 20 bp on both sides of the middle of the probe with a mapping quality score (MAPQ) >15 to be considered as supporting the corresponding allele. Genotypes were inferred from the minor allele read ratio, calculated as the ratio between the number of reads matching the less-supported allele and the total informative reads for each variant and individual, corrected by the number of available probes. Homozygotes were defined by a minor allele ratio <0.05 or >0.95 and a probability of being undetected heterozygotes of <0.03, according to the binomial probability of the observed reads based on the fraction of probes interrogating it (which for a typical inversion with two probes for each orientation corresponds to a minimum of five *O1* or *O2* supporting reads) (Puig et al. 2015b; Vicente-Salvador et al. 2017). Heterozygotes were defined by a minor allele ratio of 0.15-0.85 and at least two reads supporting each allele. All other cases were considered not reliable and were not genotyped (Figure S5).

However, six InvDups exhibited >5% of unreliable genotypes (Figure S5), caused by a continuous range of minor allele read ratio values, without a clear distinction between homozygotes and heterozygotes. This was probably due to a spurious mapping of some of the probes between the alternative alleles, resulting in a significant bias of the expected minor read ratios. Therefore, for these variants more stringent genotype filtering criteria were applied, consisting on the presence of a minimum of 10 reads supporting just one allele for homozygotes (corresponding to a minor read ratio of 0 or 1) and minor read ratios ranging from 0.3 and 0.7 for heterozygotes.

Finally, all the experimental and/or bioinformatic genotypes of 95 to 3,202 individuals from diverse 1KGP populations generated in this work, together with those available from previous studies (Aguado et al. 2014; Vicente-Salvador et al. 2017; Giner-Delgado et al. 2019; Puig et al. 2020; Campoy et al. 2022) were merged in a single VCF file.

### Variant frequency and population differenciation (*FST*) analyses

Whenever possible, frequency of the variants was estimated based on the BreakSeq obtained genotypes for the 3,202 1KGP-HC samples, and for the rest of inversions only PCR-based genotypes were used instead. Related samples according to 1KGP information were excluded in both cases. For variants with known ancestral alleles, the DAF was reported, whereas for potentially recurrent NAHR inversions in which the ancestral orientation is not clear the MAF was estimated (Table S4). Frequencies were calculated for the global population and each superpopulation separately: African (AFR), admixed American (AMR), East Asian (EAS), European (EUR) and South Asian (SAS). All variants are considered biallelic except for inversion HsInv1122, in which there is a biallelic deletion (*Del*) that removes all the inverted region, and it can have three possible alleles (*O1*, *O2* and *Del*) (Vicente-Salvador et al. 2017).

To assess whether our set of inversions and InvDups showed any frequency bias, we compared their frequency distribution to a set of neutral SNPs across the genome. Neutral SNPs with ancestral information were obtained from the 1KGP-HC data (Byrska-Bishop et al. 2022), after removing those with high conservation in mammals (GERP score >2) (Davydov et al. 2010) or a predicted high functional impact according to the Ensembl Variant Effect Predictor (VEP) (v. 110.1) (McLaren et al. 2016). Both inversion-like variants and SNPs with global MAF <0.05 were filtered out to avoid problems with missing rare variants in our dataset and Chr. Y variants were excluded. As the frequency distribution of different types of inversions and InvDups might not be comparable, due for example to NAHR inversion recurrent events (Giner-Delgado et al. 2019; Puig et al. 2020), the analysis was performed on each type of variant separately. Since in most cases the ancestral allele is not known, the MAF was used for NAHR inversions, while the DAF was used for NH inversions and InvDups. In addition, frequency was estimated for the three superpopulations with more genotyped samples, using in the calculation only individuals from YRI (AFR), CHB and JPT (EAS) and CEU (EUR) populations, for which both PCR-based and BreakSeq genotypes were generated. For each superpopulation, we compared the mean frequencies of the different set of inversion-like variants with that of 10,000 random samples of the same number of SNPs as our variants. Empirical *P*-values were estimated as the fraction of SNP samples with mean frequency values equal or more extreme than the one observed (Giner-Delgado et al. 2019).

The fixation index (*F_ST_*) (Holsinger and Weir 2009) was used to study variant frequency differences between population groups, both globally and between pairs of superpopulations, which could be an indication of the action of selection. As previously described (Giner-Delgado et al. 2019; Puig et al. 2020), inversion and InvDup *F_ST_* values were compared with empirical null distributions of 1000GP-HC SNPs with similar frequency and chromosome type (autosomes or Chr. X) to the variants in our dataset. To build the null distribution, biallelic SNPs located in easy-to-sequence regions according to the Genome in a Bottle (GIAB) Consortium (The global alliance for genomics and health benchmarking team, 2019) were selected, and their global MAF was categorized into frequency bins ranging from 0 to 0.50 with 0.01 increases. Then, for each variant in our dataset, 10,000 autosomal or 1,000 Chr. X SNPs in the same frequency bin were randomly selected without replacement. We differentiated between autosomes and Chr. X because *F_ST_* estimates in Chr. X tend to vary from the rest of chromosomes (Holsinger and Weir 2009). Finally, the F_ST_ index between all and each pair of superpopulations was calculated for variants and SNPs with vcftools (v. 0.1.17) (--weir-fst-pop option) (Danecek et al. 2011), using always just the same populations in which the inversion-like variant was genotyped. Since *F_ST_* values range between 0 and 1, negative values were converted to 0. *F_ST_* values of our variants in the top or bottom 5% of the SNP distribution were considered significant.

### Linkage disequilibrium (LD) and imputation

Pairwise LD (*r^2^*) between the available PCR-based and BreakSeq genotypes of 194 inversion-related variants (excluding a singleton inversion and two located in Chr. Y and including the two deletions) and neighboring biallelic SNPs, small indels and ESVs from the 1KGP-HC dataset (Byrska-Bishop et al. 2022) located up to 500 kb at each side of the target variant was calculated with PLINK v.1.90b7.2 (Purcell et al. 2007). For variants genotyped in a reduced number of samples by PCR-based methods, tag SNPs were strictly defined by a global perfect LD (*r^2^*= 1) across individuals of all the analyzed populations. For variants genotyped in the much larger 1KGP-HC dataset by BreakSeq, we extended the tag SNP definition to those in almost perfect LD (*r^2^* > 0.975), to allow for a small number of discrepancies between the variants and tag SNPs. Tag SNPs showing the maximum *r^2^* with the inversion-related variants were used to infer their genotypes in additional EUR and AFR 1KGP samples not genotyped and in other datasets with functional data, such as GTEx, according to the imputation decision tree shown in Figure 11. When there were several SNPs with the same maximum *r^2^*, we calculated an LD matrix of the most linked tag SNPs in the target samples to detect blocks of SNPs with the highest LD between them. Then, the genotype of each sample was established based on the consensus of the tag SNPs in the block closest to the inversion-related variant. Variant genotypes not supported by at least 75% of the tag SNPs were considered not reliable and were discarded. In addition, for inversion-like variants with more than 10 discrepancies between BreakSeq and tag SNP genotypes in 1KGP-HC data, imputation was used for GTEx samples (see below).

**Figure 11.**
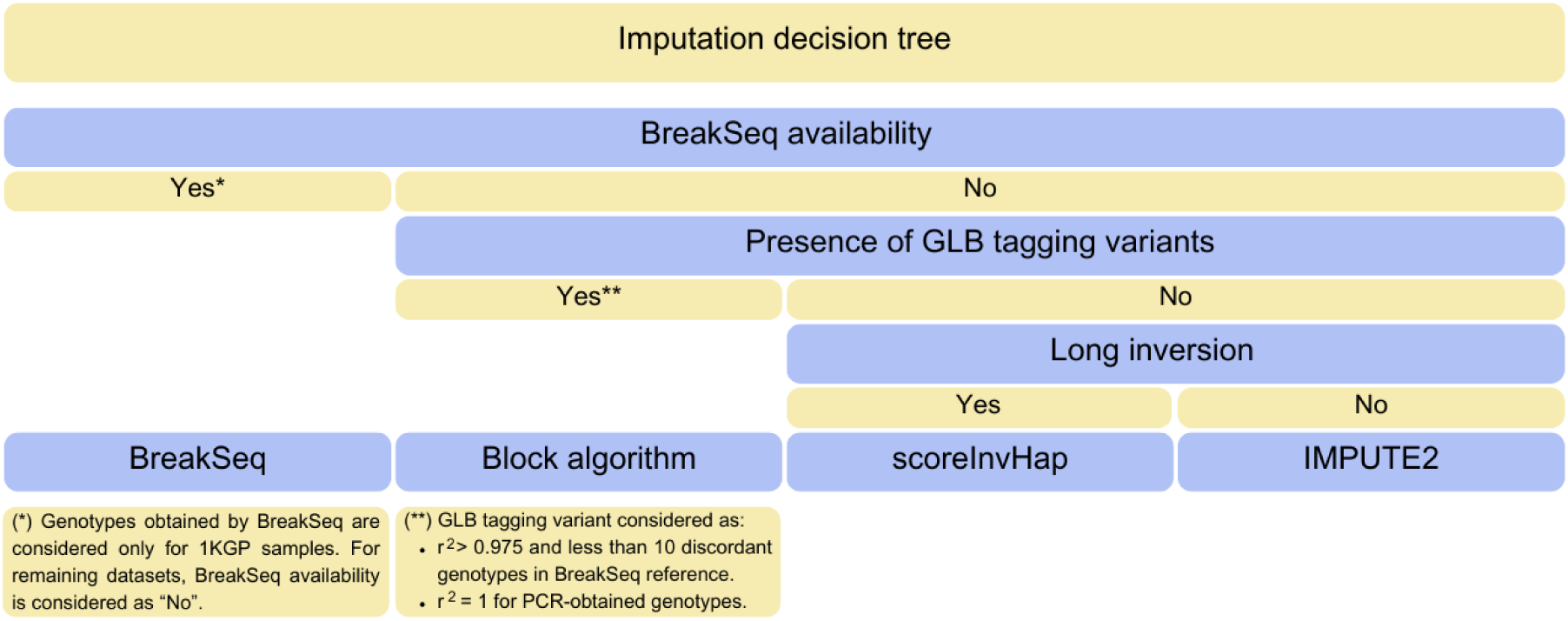
Decision tree for obtention of genotypes of inversion-related variants in additional samples with functional data. The imputation method for each variant was determined by three conditions: (1) the availability of BreakSeq genotypes for most 1KGP samples; (2) the presence of perfect (*r*² = 1) or almost perfect (*r*² ≥ 0.975 and less than 10 discordant genotypes compared to BreakSeq); and (3) the size of the inversion. Basically, BreakSeq genotypes were used whenever possible, and otherwise global tag SNPs were used if available instead. Finally, IMPUTE2 was used for the remaining variants, with the exception of two long inversions that were imputed with scoreInvHap.

In the case of the triallelic HsInv1122 variant (*O1*/*O2*/Del), for which perfect tag SNPs for both the inversion and deletion had been previously found (Vicente-Salvador et al. 2017), the inversion tag SNPs were filtered out from the final 1KGP-HC and GTEx v8 VCFs due to an excess of missing genotypes caused by the deletion. Thus, the presence of tag SNPs in a larger number of samples was confirmed using non-filtered VCFs from the 1KGP-HC (http://ftp.1000genomes.ebi.ac.uk/vol1/ftp/data_collections/1000G_2504_high_coverage/working/20200921_3202_NYGC_GATK_preQC/CCDG_14151_B01_GRM_WGS_2020-08-05_chr3.recalibrated_variants.vcf.gz) (Table S6), and the inversion genotype in GTEx v8 samples was inferred from available VCF files prior to quality control (GTEx_Analysis_2017- 06-05_v8_WholeGenomeSeq_838Indiv_AllVar_QC_metrics.vcf.gz). Conversion between hg19 and hg38 SNPs was done from the rs codes available in the file https://ftp.ncbi.nih.gov/snp/organisms/human_9606_b151_GRCh38p7/VCF/00-All.vcf.gz. For HsInv1122 genotype reconstruction, first we determined the deletion genotype and then the inversion genotype of the non-deleted alleles using the tag SNP blocks as described before. No inconsistent genotypes between the inversion and deletion (such as heterozygotes for both variants or deletion homozygotes with inversion genotypes) were found.

For inversion-related variants without tag SNPs, genotypes in additional samples with functional data were mainly determined by imputation with IMPUTE2 v.2.3.2 (Howie et al. 2009) (Figure 11), which has been recently shown to perform well compared to other imputation algorithms (Yakymenko et al. 2026). For that, first, reference panels were built by merging the available PCR and BreakSeq genotypes for our variants and all the small variants (excluding ESVs) in the final 1KGP-HC VCF (https://ftp.1000genomes.ebi.ac.uk/vol1/ftp/data_collections/1000G_2504_high_coverage/working/20220422_3202_phased_SNV_INDEL_SV/). Both reference and imputation panels were prepared by filtering out singletons and selecting only biallelic SNPs and small indels located within the inversion-related variants and the flanking 200 kb at each side, with the imputed variant placed in the middle position between its internal breakpoints. Population-specific panels were used to impute samples of the corresponding populations, including AFR and EUR for the Geuvadis and EUR for the GTEx datasets. The only exception was HsInv0822, for which 1KGP Phase 3 and GTEx v7 data were used to generate the imputation and reference panels, due to improved imputability caused by changes in this region between the hg19 and hg38 genome assemblies. In addition, we adapted the imputation strategy to Chr. X variants by coding hemizygote males as homozygous females. To determine imputation accuracy, a leave-one-out (for PCR-based genotypes) or leave-part-out (for BreakSeq genotypes) imputation of 1 or 5%, respectively, of the reference panel was performed for all the samples and the *r*^2^ between the real and imputed genotypes was calculated. Imputation of the variants was considered reliable if *r*^2^ ≥ 0.8, low quality if *r*^2^ = 0.6- 0.8, and non-imputable when *r*^2^ < 0.6. To improve imputation accuracy, unreliable imputed genotypes were filtered out by the posterior genotype probability (GP) score. By default, we kept only genotypes with GP ≥ 0.8, except in a few cases in which a stricter filtering of GP ≥ 0.9 resulted in a clear imputation improvement (such as going from non-imputable to imputable or low quality to reliable) (Table S7). However, if more than 50% of the original samples were removed, the variant was considered not imputable to avoid possible biases caused by a limited number of easy-to-impute haplotypes.

The long inversions HsInv0290 and HsInv0501 were imputed with scoreInvHap v.1.13.3 (Ruiz-Arenas et al. 2019), because it achieved a good imputation accuracy in some populations (Table S7). Imputation was done following scoreInvHap’s Bioconductor documentation and input files for AFR and EUR populations were created from the available PCR-based genotypes for each inversion. Imputation accuracy was evaluated directly by the *r*^2^ between experimental and imputed genotypes and the same criteria as before was used to determine whether the inversions were imputable, although in this case no genotypes were discarded base don GP values.

### Gene expression data

To determine the effect of inversion-related variants on gene expression levels we took advantage of the available GTEx v8 (GTEx Consortium 2020) and Geuvadis (Lappalainen et al. 2013) datasets. For GTEx, we retrieved gene-level quantifications in Transcripts Per Million (TPM) of samples with a EUR ancestry (58-504 depending on tissue) from the GTEx project webpage (https://gtexportal.org/home/) for all 45 tissues, except bladder and cervix due the low number of samples, and cerebellar-hemisphere that is a duplicate of cerebellum, plus two cell lines (Table S13). A PCA was performed on trimmed genotype data (one variant with MAF > 0.05 every 50 kb) to obtain the principal components that reflect the population membership and stratification. Since the variation of RNA-seq levels can be due to technical or biological causes, a set of covariates was applied to correct gene expression by potential confounders, while retaining biological variation. To do this, the same PCA approach was applied to the expression levels to capture technical confounding factors. In this case, the number of components used as experimental covariates was determined on the basis of the number of samples per tissue, adding 5 components every 50 samples. Then, gender, five genotyping principal components and a variable number of technical covariates were included in the model using QTLtools (v 1.3.1) (Delaneau et al. 2017).

The Geuvadis project includes gene expression data of LCLs of 358 EUR individuals from four different populations (CEU, FIN, GBR and TSI) and 87 AFR individuals (YRI) from the 1KGP (Table S13). RNA-seq reads (EMBL-EBI ArrayExpress experiment EGEUV-1) were aligned against the human reference genome GRCh38.p10 (excluding patches and alternative haplotypes) with STAR v2.4.2a (Dobin et al. 2013). Gene expression levels were quantified as RPKM based on the GENCODE v26 annotations (Harrow et al. 2012) and later normalized as described for GTEx expression data.

### DNA methylation and other epigenetic marks

DNA methylation levels from Illumina EPIC methylation arrays were downloaded from the GEO accession code GSE213478, which forms part of the Enhancing GTEx (eGTEx) dataset (Oliva et al. 2023). Data for nine different tissues was available with 33-142 samples per tissue: breast mammary tissue, colon transverse, kidney cortex, lung, skeletal muscle, ovary, prostate, testis and whole blood (Table S13). Methylation data was corrected using the same procedure as for the expression datasets.

In addition, we analyzed the levels of three well-studied histone modification marks (H3K27ac, H3K4me1 and H3K4me3) measured by ChIP-Seq across LCLs from 145 1KGP European individuals, which were also summarized in chromatin regulatory domains (CRDs) that are a finer chromatin organization inside TADs characterized by correlated histone modification changes that provide information on chromatin activity (Delaneau et al. 2019) (Table S11). In this case, coordinates of the different signals were converted from hg19 assembly to hg38 employing the liftOver tool from the UCSC genome browser (Hinrichs et al. 2006), and we tested the CRDs regions and the levels of the different histone modification marks separately.

Finally, as another functionally relevant epigenetic mark, we analyzed DNase I hypersensitive sites from 59 1KGP YRI LCLs (Degner et al. 2012), which measures chromatin accessibility (Table S13). Normalized levels from DNase I hypersensitive sites sequencing (DNase-seq) were downloaded from GEO accession number GSE31388, and DNase-seq levels in 100 bp windows were converted from assembly hg18 to hg19 and hg38 using the liftOver tool (Hinrichs et al. 2006).

### Cis QTL mapping

For the quantification of the effect of inversion related variants on the different types of molecular traits, only those genotyped directly by PCR-based methods or BreakSeq and inferred by tag SNPs or reliable imputation (*r*² ≥ 0.8) were used. For each variant, we created a VCF centered in the variant coordinates plus 1 Mb at each side that included only biallelic and non-singleton variants from the samples in the corresponding dataset. The genotypes of our variants were introduced in each breakpoint position, and in other internal positions if the distance between the breakpoints was greater than 1 Mb. Additionally, for 14-18 NAHR inversions without tag SNPs that were not genotyped with BreakSeq or reliably imputed in the 1KGP-HC dataset, available PCR-based genotypes were included in the analysis of Geuvadis LCL gene-expression levels, histone modifications and CRDs. The number of inversions and InvDups tested varied depending on their genotype quality, their frequency, and the number of samples in each dataset or tissue. To ensure the reliability of the results, only variants with MAF ≥ 0.01 in the appropriate samples selected according to their ancestry (EUR or AFR) were analyzed (Table S13).

Variants acting as quantitative trait loci (QTL) in *cis* were identified by testing their association with gene expression or other epigenetics marks from the datasets described above. To quantify the relative contribution of the variants to molecular changes, a joint *cis*-QTL analysis was conducted, including all neighboring small variants (SNPs, indels) detected in each dataset, along with the reliable genotypes of inversion-like variants. For expression levels of genes and different epigenetic marks (DNA methylation, histone modifications and CRDs), only those located within a 1 Mb window (considering the transcription start site (TSS) for genes) of at least one autosomal or Chr. X inversion or InvDup were included in the analysis. For DNaseI hypersensitivity data, due to the large number of windows, associations were restricted to 5 kb regions on either side including an inversion or InvDup. Molecular measurements were transformed into standard normal distributions, followed by linear regression using QTLtools software (v 1.3.1) (Delaneau et al. 2017).

Multiple-testing correction of *cis*-QTL results was done at two different levels (Aguet et al. 2023). First, 1,000 genotype-phenotype permutations were performed to correct for the multiple variants per molecular trait, accounting for the LD among variants. Second, to remove potential false positives due to the high number of molecular traits across the genome, for each GTEx tissue and each molecular dataset, a multiple-testing correction including all the tested variants was carried out using the Benjamini-Hochberg method at 5% FDR with the qvalue package (v.2.18.0) (Storey 2002) in R (v.4.3.1) (R Core Team 2021). Moreover, to ensure the reliability of the *cis*-QTL results, only inversions or InvDups acting as lead markers or in high LD with the top lead QTLs (*r*^2^ ≥ 0.8) were reported.

### Detailed analysis of inversion-related variant effects

To evaluate the potential mutational effects of inversion-like variants on genes, we compared their breakpoint coordinates with the GENCODE comprehensive annotation dataset v26, taking into account gene isoforms with a transcript support level of 3 or higher, single-exon genes not labeled as "problem", and pseudogenes (Harrow et al. 2012). Variants were classified into the following categories: "Intronic", when both breakpoints were located entirely within an intron; "Gene/Transcript/Exon disruption", when one breakpoint was entirely included within a gene, transcript or exon; "Whole gene/exon inversion", when the entire gene or exon was contained within the inverted region; "Breakpoint overlapping gene", when genes were fully comprised within the IRs at the breakpoints; and "Intergenic", when a variant did not meet any of the above criteria.

The distance between lead inversion-like variants and the TSS of the possible gene they affect was calculated as 0 bp when one of the variant breakpoints or the whole variant was included within the gene. For all other cases, we calculated the distance between the gene TSS and the closest of the variant four breakpoints coordinates. This approach led to some values exceeding the 1 Mb threshold used to include different variants in the QTL mapping analysis, but it was a more accurate way of assessing the shortest distance between genes and inversions.

To study in detail the possible molecular consequences candidate inversions that affect directly gene sequences, such as HsInv0030 and HsInv1117, we first modified the hg38 assembly by reversing *in silico* the sequence between the inversion breakpoints (HsInv0030, chr16:75205642-75223319; HsInv1117, chr17:18622233-18823774). We used the STAR 2- pass pipeline (https://docs.gdc.cancer.gov/Encyclopedia/pages/STAR_2-Pass_Genome/) to map the RNA-Seq reads extracted from the GTEx project BAM files from available O1 and O2 homozygotes against the genome with the inverted conformation, and only uniquely mapped reads were selected. Tissues assayed were pancreas for *CTRB1* and *CTRB2* (HsInv0030), and adipose subcutaneous and testis for *AKR1C1* and *AKR1C2* (HsInv1117), where the genes affected by the inversions were highly expressed. For HsInv1117, the same process was repeated for Geuvadis LCL reads. RNA-seq profiles were computed separately for homozygotes of each inversion orientation, and transcript structures were reconstructed with Cufflinks default parameters by merging all reads from each genotype. RNA-seq profiles were visualized on the Integrative Genomics Viewer (Robinson et al. 2011).

### Association of inversion-like variants and phenotypic traits

To check if polymorphic inversion-like variants are associated with specific traits or diseases, we took advantage of the NHGRI-EBI Catalog of human genome-wide association studies NHGRI Catalog of published GWAS (http://www.ebi.ac.uk/gwas/) [release 2023-04-07, v1.0], which stores a curated collection of the most significant SNPs from each independent locus highly associated to a particular phenotype (MacArthur et al. 2017). First, we investigated if there was enrichment in the number of trait-associated signals in the variants and flanking regions (± 20 kb) compared to what should be expected by chance. To do so, we selected 1KGP-HC SNPs within these regions and crossed them with GWAS Catalog signals with 1KGP-HC variants. GWAS SNPs in high LD (*r*^2^ ≥ 0.8) and associated exactly with the same phenotype were grouped together to obtain a non-redundant set. Next, we carried out a permutation strategy to generate 100 random genomic regions as a null model for each variant and tested observed versus expected GWAS hits for the whole set of variants. To avoid biasing the results, the Chr. Y was excluded of this analysis and permuted regions could not overlap genome gaps. We also extended the analysis to explore which individual inversions were more significantly enriched in GWAS signals. Therefore, we repeated the same procedure, but using a one-tailed permutation test for each inversion to deal with inversions including zero GWAS signals.

Lastly, we crossed GWAS Catalog significant variants with those SNPs in high LD with the inversion-like variants (*r^2^* ≥ 0.8), based in PCR or BreakSeq genotypes. Population ancestry of the study where GWAS signals were reported was taken into account to select the appropriate LD value between our variants and the SNPs. For each population, the LD of the corresponding super-population (EUR, AFR, AMR, EAS and SAS) was selected (e.g. EUR for British or Spanish), whereas the global LD was selected for populations from different continents.

## Data Availability

All data produced in the present work are contained in the manuscript.

## Acknowledgements

We thank Clara Vizuete and former members of the Comparative and Functional genomics lab for their help with human inversion validation and genotyping. BreakSeq genotyping was performed in collaboration with the Port d’Informació Científica (PIC) data center. PIC is maintained through a collaboration agreement between the Institut de Física d’Altes Energies (IFAE) and the Centro de Investigaciones Energéticas, Medioambientales y Tecnológicas (CIEMAT). This work was supported by research grants BFU2016-77244-R, PID2019- 107836RB-I00 and PID2022-137615OB-I00 funded by the Agencia Estatal de Investigación of the Ministerio de Ciencia, Innovación y Universidades (MICIU/AEI/10.13039/501100011033, Spain) and the European Regional Development Fund (ERDF, EU), ERC Proof of Concept Grant 755027 (IN2DIAG) from the European Research Council under the European Union Seventh Research Framework Programme (FP7), and 2021 SGR 00526 support to research groups from the Departament de Recerca i Universitats (Generalitat de Catalunya, Spain) to MC, a La Caixa Doctoral fellowship to JLJ, Predoctoral Contracts BES-2017-082018 to EC and PRE-2020-092440 to IY funded by the Agencia Estatal de Investigación of the Ministerio de Ciencia, Innovación y Universidades (MICIU/AEI/10.13039/501100011033, Spain), and the predoctoral program AGAUR-FI Joan Oró grants 2019 FI-B 01321 to RGG and 2021FI-B 00240 to RMP funded by the Departament de Recerca i Universitats (Generalitat de Catalunya, Spain), as well as the European Social Plus Fund,). MP is a Serra Húnter Fellow.

## Notes

### Competing Interest Statement

The authors have declared no competing interest.

### Author Declarations

The Research Ethics Committee (CERec) of the Universitat Autonoma de Barcelona (CEEAH 3550) and the Hospital del Mar Clinical Research Ethics Committee (CEIm, 2023/11107/I,) gave ethical approval for this work.

