## Supplemental Information for "Towards a complete characterization of common human polymorphic inversions and their functional effects"

* Equally contributing authors.

[†] Corresponding author.

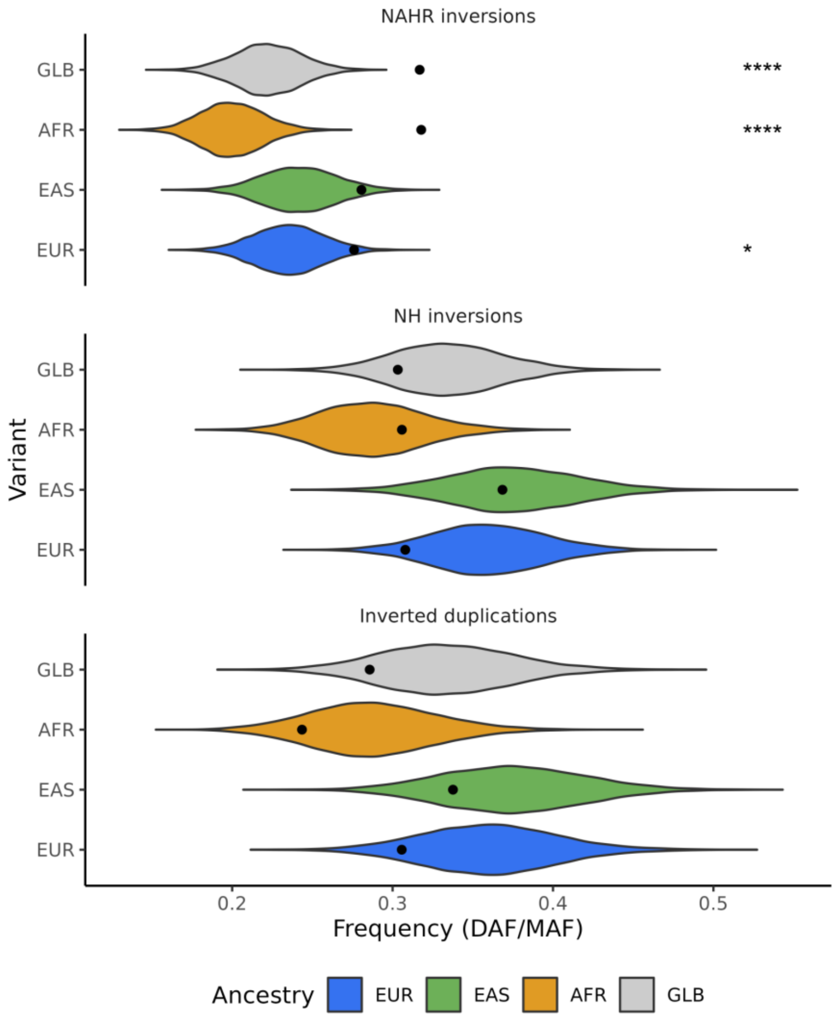


**Figure S1.** Frequency comparison between different types of inversion-like variants and SNPs across populations. Observed average frequency of NAHR and NH inversions and inverted duplication in each population and all the populations together (GLB) (black dots) compared with that expected from a null distribution of 10,000 SNP samples (colored violin plots). Minor allele frequency (MAF) is shown for NAHR inversions and derived allele frequency (DAF) for other variants. The *P* values represent 1 minus the percentile of the observed average frequency in the SNP distribution.

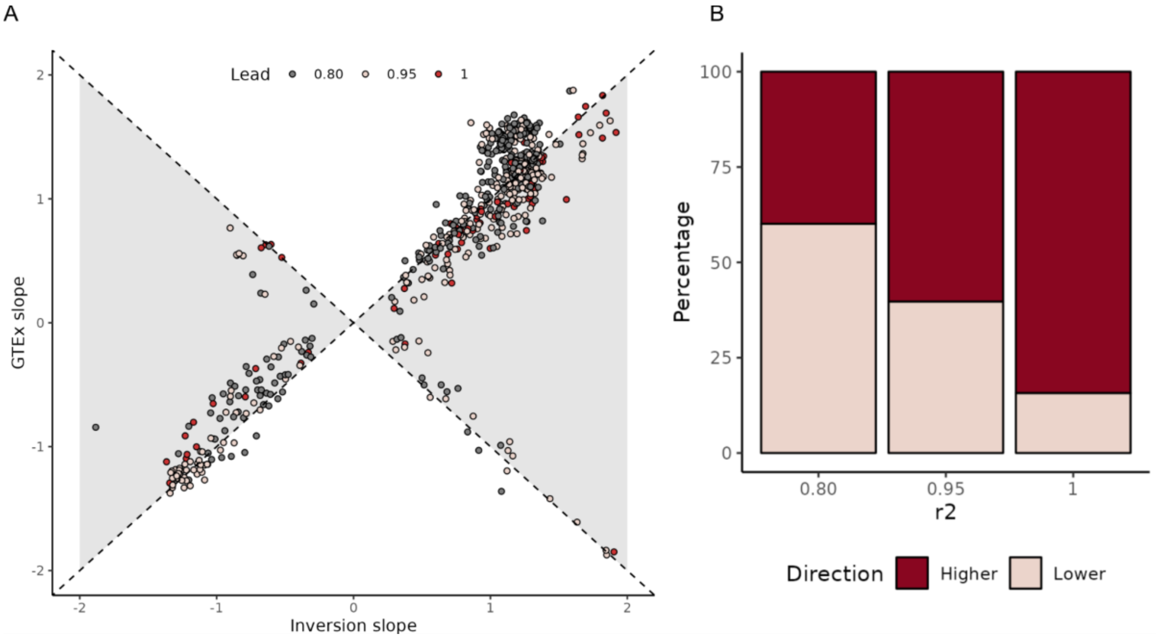


**Figure S2. Comparison of the effects on gene expression between inversion-like variants and SNPs from GTEx. A.** Gene expression effect (represented as the association slope) for inversion-like eQTL variants reported in this study and eQTL SNPs for the same gene and tissue previously identified in the GTEx catalog, with the color of the dots indicating the LD of inversions and InvDups with the lead variants ($r^2 \geq 0.8$). Gray-shaded regions represent associations where inversion-like variants exhibit larger effects compared to GTEx SNPs. **B.** Proportion of inversion-like variants showing effect sizes smaller (pink) or larger (dark red) than the SNPs reported in GTEx for the corresponding gene and tissue categorized according to the LD ($r^2$) between the lead and inversion-like variant. The proportion of inversion-like variants with a higher absolute effect size increases as the LD with the lead variant is stronger ($r^2 \geq 0.95$).

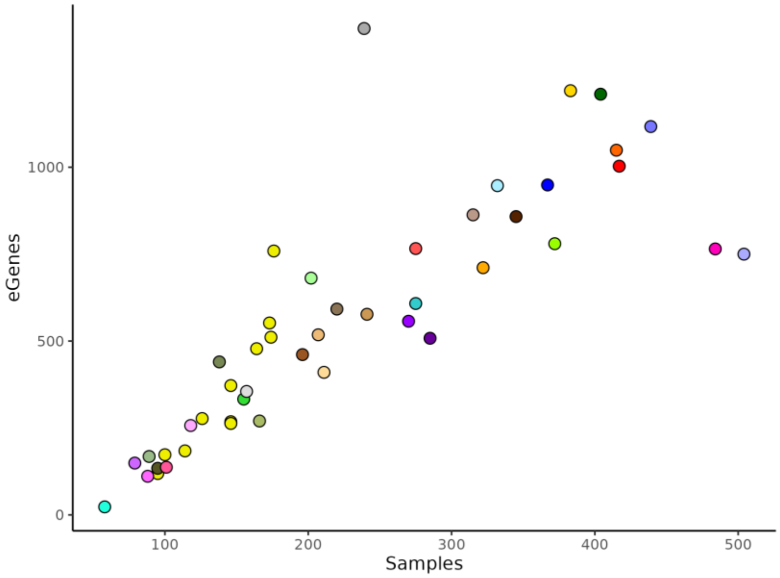


**Figure S3.** Correlation between the number of unique genes associated with eQTLs (eGenes) identified in our analysis and the number of available samples in each tissue in the GTEx expression dataset (Spearman's rank correlation coefficient = 0.92, $P < 2.2 \times 10^{-16}$). Colors represent different types of tissues.

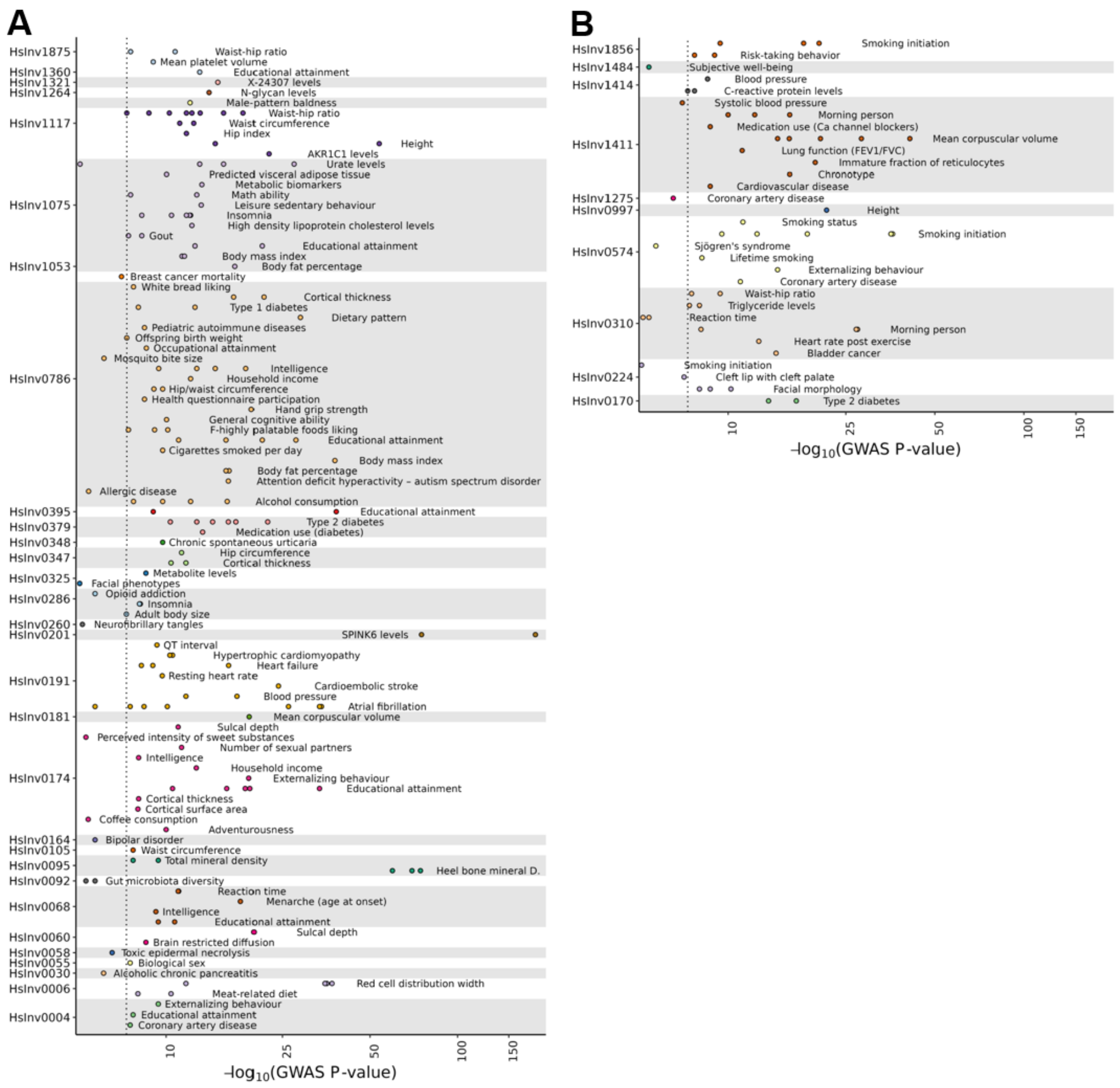


**Figure S4.** Summary of phenotypic associations for the inversions (**A**) and inverted duplications (**B**) included in the dataset. Colored dots indicate the GWAS catalog signals ($P < 0.05$) in high LD ($r^2 \geq 0.8$) in the closest studied population with the different variants (Y axis). X axis shows the minus base-10 logarithm of the reported association $P$ value, with the genome wide threshold of $P < 5 \times 10^{-8}$ indicated by a vertical dotted line. 17q21.31 and 8p23.1 inversions that had multiple phenotypic associations and had been analyzed with a previous GWAS catalog version (Campoy et al. 2022) were excluded from the representation.

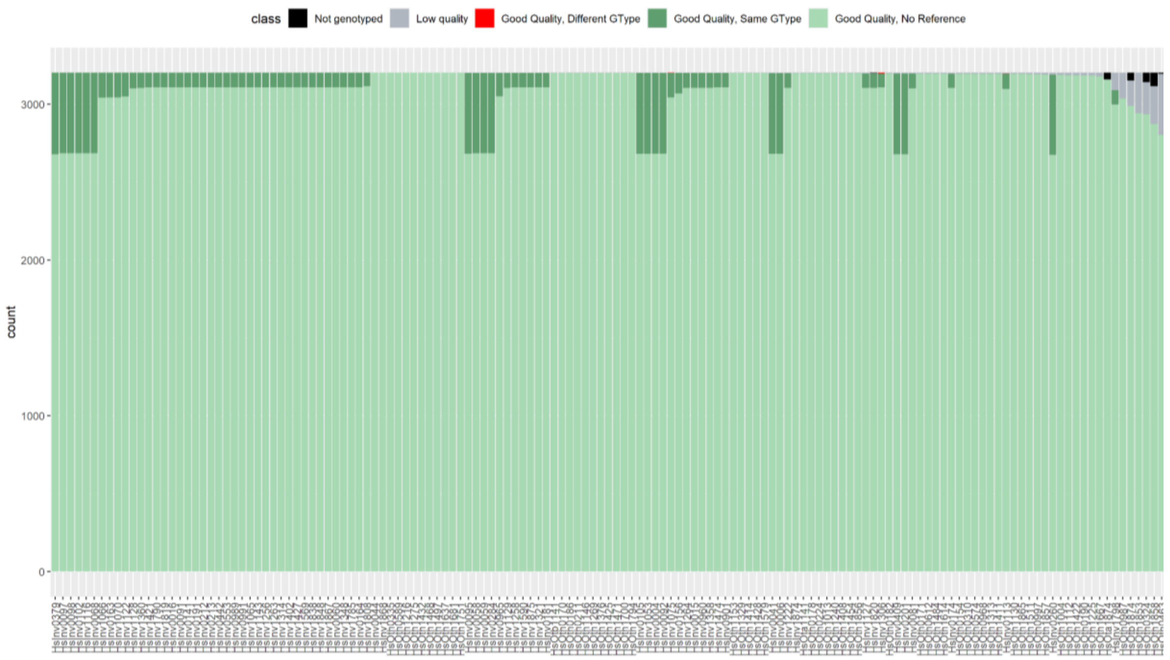


**Figure S5.** Summary of BreakSeq genotyping results for 143 inversion-related variants on the 3202 samples from the 1000 Genomes High Coverage data. Variants are sorted by BreakSeq performance along the X-axis, while the Y-axis represents for each variant the number of accepted genotypes (light green) and the results of the comparison with available experimentally validated genotypes of 77 inversions in 87 to 523 individuals (with those concordant and discordant with experimental genotypes in dark green or red, respectively). Low reliable genotypes according to standard BreakSeq filtering criteria are shown in gray and those samples without any reads in black.
